# Identifying the viral and epidemiological factors behind the apparent global extinction of influenza B/Yamagata

**DOI:** 10.64898/2026.07.22.26358639

**Authors:** Maria A. Gutierrez, Pejman Rohani

## Abstract

Until 2020, two lineages of the influenza B virus had co-circulated globally. The COVID-19 pandemic led to a near-absence of influenza infections. While B/Victoria reemerged in late 2021, there have been no reports of B/Yamagata since the pandemic. To investigate which epidemiological and immunological factors were primarily responsible for the extinction of B/Yamagata, we developed a global model for the two influenza B lineages. To mimic the transmission impacts of the pandemic, we implemented a transient reduction in contacts and identified parameter values that recapitulated viral coexistence dynamic before the pandemic and the qualitative post-pandemic outcomes of B/Victoria (reemergence in late 2021) and B/Yamagata (extinction). Our results suggest that, rather than immunological or evolutionary mechanisms, the extinction of B/Yamagata was mainly driven by its lower basic reproduction number making the virus particularly vulnerable during the early phase of the pandemic. Stochastic simulations of our best-fitting model suggest that B/Victoria was also close to extinction during this period. We investigate the model to assess the feasibility of B/Victoria eradication through vaccination and the potential for a sustained re-emergence of B/Yamagata in the 2026-27 flu season, thus highlighting important considerations for biosafety.

**Significance:** For decades, the Yamagata and Victoria lineages of the influenza B virus co-circulated globally, causing millions of infections in humans. Only Victoria reemerged after the COVID-19 pandemic. By combining mechanistic transmission modeling with worldwide influenza surveillance data, we show that Yamagata’s extinction is most consistent with a lower intrinsic transmissibility than that of the surviving Victoria lineage. Contrary to other proposed explanations, differences in cross-immunity and immune escape appear insufficient to explain the observed outcome. Our analyses further indicate that Victoria narrowly avoided extinction during the COVID-19 disruption, but now transmission of Yamagata would be sustained if the lineage were reintroduced. These results inform both influenza eradication efforts and biosafety policy.

---

Influenza B virus was first reported in 1940 (1, 2). In the 1970s, influenza B diverged into two antigenically distinct lineages: ‘B/Victoria/2/87-like’ and ‘B/Yamagata/16/88-like’ (3, 4), henceforth referred to as Victoria and Yamagata. Both lineages co-circulated globally since at least 2001 until 2020 (5) and account for ~20% of the estimated 650 million yearly cases of seasonal influenza, leading to substantial public health and economic burdens (6). Historically, the epidemics of influenza B were subject to strong seasonality in temperate regions, producing yearly winter epidemics (6) (Fig. 1). Year-round transmission in tropical regions seeded epidemics outside of the tropics (7, 8). These global circulation patterns were also shaped by antigenic evolution (7), which was substantial in both Yamagata and Victoria (9). Yamagata was thought to have evolved more slowly than Victoria prior to 2013, with stronger selection pressure on the hemagglutinin gene of Victoria (4). However, it has been proposed that selection in the neuraminidase gene of Yamagata might have increased the epidemic activity of Yamagata from 2015 to 2019 (4).

**Fig. 1.**
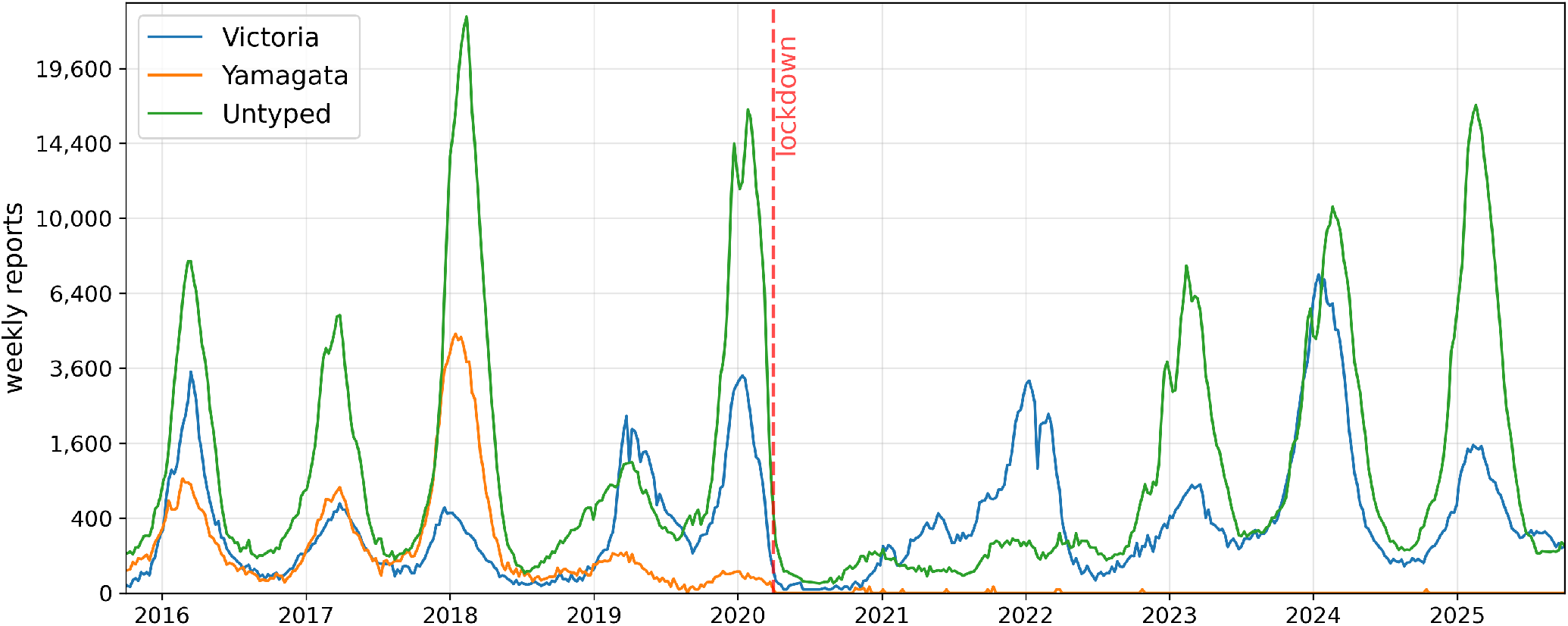
FluNet data for influenza B infections in Northern Hemisphere Temperate and Sub-tropical regions, from October 1st, 2015 until September 30th, 2025. Similar pre- and post-pandemic patterns for influenza B are observed in the Tropics and Southern Hemisphere (Fig. S1).

The COVID-19 pandemic (caused by SARS-CoV-2 and henceforth referred to as ‘the pandemic’) substantially disrupted the transmission of many airborne diseases (10, 11), including seasonal influenza viruses (10, 12, 13). Coinciding with the roll out of social distancing measures in response to the pandemic (14, 15), there were no outbreaks of either influenza B lineage in the 2020-21 winter season of the Northern Hemisphere (16). While Victoria reemerged in late 2021 and has been circulating ever since, Yamagata has most likely gone extinct (17). It is important to identify the phenotypic differences responsible for the contrasting post-pandemic outcomes for Victoria and Yamagata. Putative hypotheses include (i) different rates of antigenic drift, which effectively modify the average duration of protection induced by infection and vaccination (4, 7, 18); (ii) asymmetric cross-immunity, whereby infection with Victoria protects against Yamagata but the converse is not true (19); and (iii) differential reproduction number, with Victoria estimated to be more transmissible than Yamagata (20). Except for (18), no work to date has tested these hypotheses or other potential mechanisms behind the apparent extinction of Yamagata.

Here, we investigate the factors underlying the contrasting fates of Yamagata and Victoria using a mechanistic model for the global ecology of influenza B viruses. We search through one million plausible parameter combinations, spanning a range of plausible epidemiological and immunological scenarios, and identify the conditions that recreate the observed pre- and post-pandemic patterns. Our results indicate that Yamagata’s extinction is most consistent with a lower intrinsic transmissibility than Victoria, rather than differences in cross-immunity or antigenic evolution. We then use stochastic simulations to evaluate how close Victoria was to extinction, as well as the current potential for sustained Yamagata spread upon re-introduction.

## Results

Influenza B incidence data from FluNet (21), a global web-based surveillance tool coordinated by the World Health Organization (WHO), show noisy annual epidemics of Victoria and Yamagata before the pandemic (Fig. 1). Since late 2021, yearly B/Victoria epidemics in the Northern Hemisphere have resumed, while, in contrast, Yamagata has not been detected since the spring of 2020 (with the exception of some vaccine-derived infections (17)). To understand the mechanisms underlying these dynamics, we developed a global two-lineage epidemiological model, aimed at recreating the qualitative epidemic dynamics of influenza B before and in response to the pandemic. Our model consists of 3 separate populations, linked by movement: Northern Hemisphere (NH), the tropics and Southern Hemisphere (SH). To mimic the impact of COVID-19, we include a temporary reduction in transmission (‘lockdown’), which is *c*_0_ ∈ [0, 1] at the start of the pandemic and decays exponentially at rate 1*/τ*. Given the short time-series of viral incidence and the uncertainty in key parameters (determining the reduction in transmission during lockdown, the extent of cross-immunity and the epidemiological characteristics of influenza B viruses), we opted for a feature-matching strategy (22–24) rather than standard likelihood estimation (25, 26). For this, we generated a million parameter combinations using Latin Hypercube Sampling (27) for ten model parameters (Table 1). The trajectories that each parameter set produced were then assessed for qualitative agreement with the observed epidemiology of influenza B viruses; specifically, coexistence prior to the pandemic, the re-emergence of Victoria between Oct 2021 and March 2022 and the extinction of Yamagata (Fig. 1).

**Table 1.** Distribution of model parameters after searching one million samples for matching pre- and post-pandemic influenza B patterns.

| Parameter | Bounds | **Mean** | (25th, **50th**, 75th) percentiles | Mean \|SHAP\| |
|---|---|---|---|---|
| B/Vic's basic reproduction number: $R_{0,V}$ | (1, 5) | **2.63** | (2.05, **2.54**, 3.10) | 2.0 |
| B/Yam's basic reproduction number: $R_{0,Y}$ | (1, 5) | **2.02** | (1.47, **1.86**, 2.37) | 2.0 |
| Average duration of B/Vic's immunity (years): $T_V$ | (1, 10) | **6.22** | (4.30, **6.48**, 8.34) | 1.2 |
| Average duration of B/Yam's immunity (years): $T_Y$ | (1, 10) | **6.64** | (4.97, **7.00**, 8.57) | 1.0 |
| Cross-immunity from B/Vic to B/Yam (probability of becoming immune): $\sigma_{VY}$ | (0, 1) | **0.32** | (0.11, **0.27**, 0.48) | 0.5 |
| Cross-immunity from B/Yam to B/Vic (probability of becoming immune): $\sigma_{YV}$ | (0, 1) | **0.48** | (0.24, **0.47**, 0.72) | 0.2 |
| Inter-region contact rate (relative): $m$ | (e-3, e-2) | **5.3e-3** | (3.0e-3, **5.3e-3**, 7.5e-3) | 0.3 |
| Peak seasonality in NH (days since Jan 1st): $\phi$ | (0, 100) | **47** | (24, **45**, 68) | 0.2 |
| Initial COVID-19 reduction in contacts (relative): $c_0$ | (0.2, 0.8) | **0.36** | (0.31, **0.35**, 0.40) | 3.0 |
| Lockdown timescale (days): $\tau$ | (100, 600) | **388** | (276, **407**, 509) | 1.3 |

### Yamagata extinction was likely due to lower intrinsic transmissibility

Across these one million parameter combinations, the coexistence of Victoria and Yamagata before 2020 was observed in 43% of parameter sets. The gray violin plots in Fig. S2 show how these ‘coexistence’ samples are distributed for each parameter. Among parameter samples that successfully captured pre-pandemic coexistence, only 0.90% (*i*.*e*., 0.38% of total samples) generated the observed post-pandemic outcomes. These successful samples are distributed non-uniformly across parameter space: the mean and the 25th, 50th (median), and 75th percentile values for each parameter are shown in Table 1 (the full distributions appear in Fig. S2). The distributions for the basic reproduction number of each lineage are both unimodal, with that of Yamagata (*R*_0,*Y*_) clustered around a lower value (mean = 2.02) than that of Victoria (*R*_0,*V*_; mean = 2.63). Interestingly, in these selected samples, the initial reduction *c*_0_ in contacts due to COVID-19 lockdowns is clustered around relatively low values (*<* 50%). In contrast, the observed post-pandemic outcomes are skewed around a relatively long lockdown timescale, *τ* (*>* 1 year).

To quantify the contribution of each parameter to the feature selection outcome (whether a sample matches pre- and post-pandemic patterns), we trained an XGBoost classifier (28) and computed SHAP (SHapley Additive exPlanations) values (29). The distributions of SHAP values are shown in Fig. S3 and the mean absolute SHAP values are presented in Table 1. The three parameters with the largest mean |SHAP| are: the initial reduction in contacts (*c*_0_), the reproduction number of Yamagata (*R*_0,*Y*_), and the reproduction number of Victoria (*R*_0,*V*_). This is consistent with the distributions discussed above and can be understood intuitively. High values of *c*_0_ have large negative SHAPs because, in the model, the lockdown must be mild enough to allow the reemergence of Victoria. The reemergence of Victoria in the appropriate time window requires an intermediate *R*_0,*V*_ : both low and high values of *R*_0,*V*_ lead to large negative SHAP values because they either led to Victoria extinction during the pandemic or a rebound that is much earlier than observed (Fig. S3). Finally, the extinction of Yamagata requires low *R*_0,*Y*_ values (these have the largest positive SHAP values; Fig. S3).

To understand how model parameters interact to shape post-pandemic outcomes, we projected the selected samples into four two-dimensional parameter spaces (Fig. 2). In Fig. 2A, we observe a linear bound on the basic reproduction number of Yamagata (*R*_0,*Y*_) as a function of the average duration of immunity to Yamagata (*T*_*Y*_). Below this bound, there are two explanations for why samples are not consistent with data: a relatively high *R*_0,*Y*_ implies either that (i) Yamagata reemerges (bottom right corner without dots) or that (ii) Victoria also has a relatively high reproduction number—to ensure pre-pandemic coexistence—which means Victoria reemerges early (yellow dots). The green dots in Fig. 2B represent parameter combinations where the timely reemergence of Victoria requires a reproduction number *R*_0,*V*_ that is confined to an intermediate region, relative to the average duration of immunity *T*_*V*_. An *R*_0,*V*_ that is too high (early reemergence) or too low (late reemergence or extinction) would be inconsistent with observations. A similar tradeoff between *R*_0,*V*_ and the lockdown timescale *τ* appears in Fig. 2C: if the lockdown is relatively short, Victoria would have reemerged earlier than observed; if it is relatively long, Victoria would have reemerged later or gone extinct. Finally, among the selected samples (green dots of Fig. 2D), the lockdown timescale is inversely correlated with the initial reduction in contacts, *c*_0_. A short and mild lockdown (bottom left corner) would have led to the reemergence of Yamagata, whereas a long and strict lockdown would have led to the extinction of Victoria (top right corner). It is surprising that the red dots (late Victoria reemergence) are located at generally lower values of *c*_0_ than the yellow dots (early Victoria reemergence). This means that a stricter lockdown (higher *c*_0_) favors an early recovery, *conditional* on Yamagata extinction and Victoria reemergence. For high values of *c*_0_, Victoria either reemerges too early or goes extinct.

**Fig. 2.**
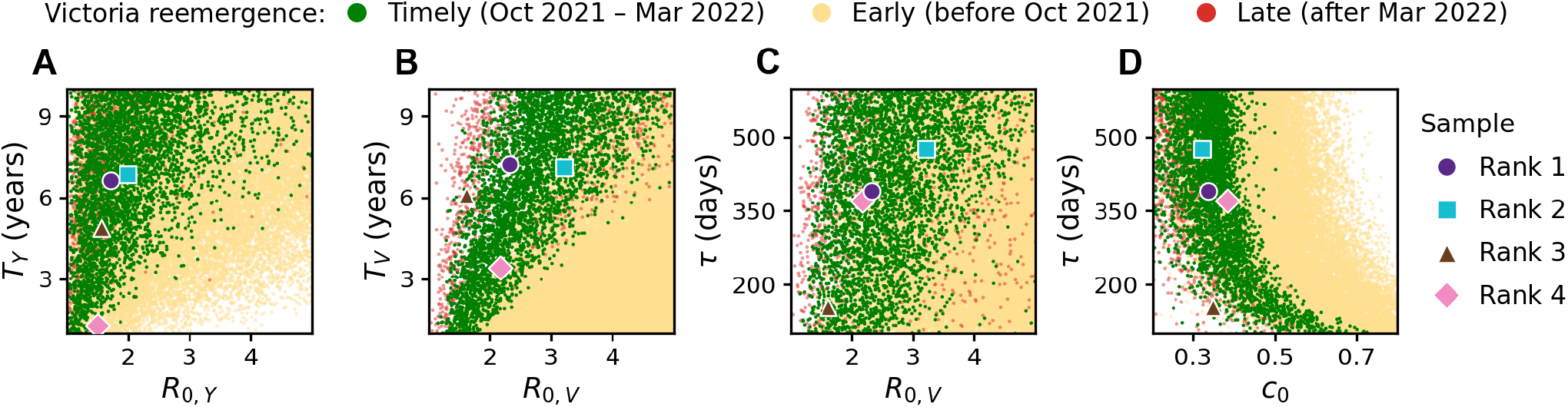
Projection into two-dimensional parameter subspaces of the samples with pre-pandemic coexistence, Yamagata extinction, and Victoria recovery. Green dots: samples with Victoria recovery within the required time window (between October 2021 and March 2022), i.e., the selected samples. Yellow dots: samples with early Victoria recovery (before October 2021). Red dots: samples with late Victoria recovery (red dots, after March 2022). Symbols mark the four best fits to the NH data (Table S1); rank 1 is the best fit shown in Fig. 3. The axes span the sampling bounds of each parameter (Table 1).

#### Victoria was also close to extinction during the lockdowns

To further ground our analysis on empirical knowns, we imposed two biological constraints based on experimental and phylogenetic data for Victoria and Yamagata. These constraints were transitive relations for the lineage-specific parameters. First, we required the average duration of immunity to be longer against Victoria than Yamagata (*T*_*V*_ *> T*_*Y*_). Here the waning of immunity implicitly represents antigenic drift. Thus, this constraint is consistent with stronger antigenic evolution of Yamagata in the 2010s (4) and Victoria providing higher within-lineage protection than Yamagata (30). Second, we required the cross-immunity elicited from Victoria against Yamagata to be stronger than the cross-immunity elicited from Yamagata against Victoria (*σ*_*V Y*_ *> σ*_*Y V*_), as established in animal challenge models (19). These two constraints were satisfied on 15% of the 3,844 parameter samples selected in the previous section (i.e., those that match the qualitative pre- and post-pandemic features of influenza B). Thus, only 0.06% of the total parameter combinations (595 out of one million) satisfied the biological constraints while recapitulating the extinction of Yamagata. These ‘filtered’ samples are distributed in parameter space similarly to the ‘selected’ samples described earlier (Figs. S2, S4, S5), except for shifts in the individual distributions of *T*_*V*_, *T*_*Y*_, *σ*_*V Y*_, *σ*_*Y V*_ (as expected from the constraints *T*_*V*_ *> T*_*Y*_ and *σ*_*V Y*_ *> σ*_*Y V*_). Only 20 of the 595 filtered samples have a higher basic reproduction number for Yamagata than Victoria (Fig. S6), which again highlights the importance of a relatively low *R*_0,*Y*_ in recreating the observed influenza B patterns.

We evaluated the 595 filtered samples against the Northern Hemisphere FluNet data (Fig. 1) using a likelihood function assuming Poisson observation errors (31). The sample with the highest likelihood (henceforth, ‘the best-fitting sample’) produced the epidemic dynamics shown in Fig. 3. Before 2020, the predicted Victoria epidemics in the Northern Hemisphere are substantially larger every other influenza season (Fig. 3A). The predicted Yamagata epidemics in the Northern Hemisphere instead follow a four-year cycle. Only in one of these four years the model predicts a higher prevalence for Yamagata than Victoria. A similar four-year pattern appears in the Southern Hemisphere (Fig. 3C). In the Tropics (Fig. 3B), the number of predicted infections slightly oscillates despite the lack of seasonality (due to the influence of the NH dynamics through global travel), but Victoria is always more prevalent than Yamagata. For this best-fitting sample, Yamagata goes extinct in the autumn of 2020 (October 10th). At its lowest prevalence, the deterministic model predicts that Victoria had 3 active infections globally. The second, third, and fourth-best fitting samples had a similarly low minimum Victoria prevalence: 18, 52, and 1 infections, respectively (Table S1). Over the 595 filtered samples (Fig. S7), the minimum Victoria prevalence had a median of 5.4 infections, interquartile range 2.4-18.5, and 95% range 1.25-405.

**Fig. 3.**
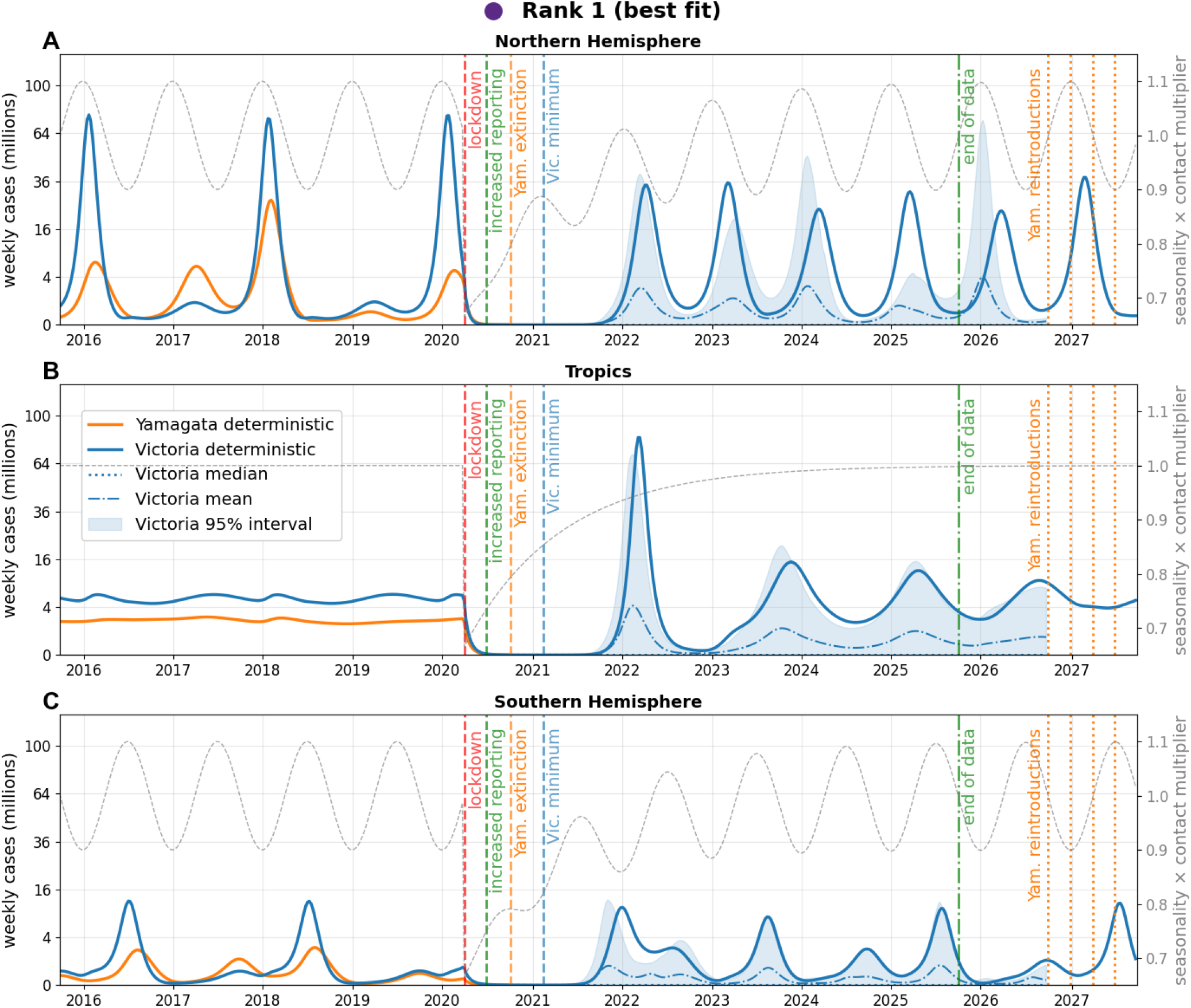
Model outcome for the biologically-filtered sample that best fits the Northern Hemisphere FluNet data (Fig. 1). The parameter values are *R*_0,*V*_ = 2.32, *R*_0,*Y*_ = 1.71, *T*_*V*_ = 7.22, *T*_*Y*_ = 6.62, *σ*_*V Y*_ = 0.31, *σ*_*Y V*_ = 0.06, *ϕ* = 3.3, *m* = 0.004, *c*_0_ = 0.34, *τ* = 390 (keys and units in Table 1). The pre- and post-pandemic reporting rates that maximize the likelihood are *ρ*_*−*_ = 0.031% and *ρ*_+_ = 0.046%. The orange dotted lines represent the hypothetical dates of Yamagata re-introduction (see Fig. 4). The blue dashed line marks the minimum global prevalence of Victoria according to the deterministic model.

The predicted low prevalence of Victoria during the early lockdown period suggests the lineage was at risk of demographic extinction, as stochastic effects have a greater impact when pathogen prevalence is low (32). Thus, we ran 105 stochastic simulations for each of the four best-fitting samples (Table S1) to examine the epidemiological dynamics of Victoria after the extinction of Yamagata. In each of these four samples, the minimum Victoria prevalence (according to the deterministic model) was reached after the extinction of Yamagata (Fig. S9). We found that, for all four samples, the 95% prediction interval of epidemic trajectories includes the extinction of Victoria (Fig. 3, Fig. S13, Fig. S15, and Fig. S17). Victoria went extinct in 93%, 60%, 39% and 97% of the stochastic runs, respectively (Table S1).

#### Yamagata would be able to spread in the current landscape of population immunity

To evaluate the reemergence risk of Yamagata (should it be reintroduced), we used the deterministic model to estimate the current population immunity against Yamagata. Waning of immunity and population turnover decrease the number of immune individuals, but cross-immunity from Victoria infections could theoretically outweigh these reductions. To measure the overall balance of these effects, we ran the model through the 2026-27 winter season and calculated *R*_*e*_, the *effective* reproduction number of Yamagata in the NH region (Equation 6). An effective reproduction number above one means that sustained transmission is possible (32). For the parameter combination which best fits the data (Fig. 3), *R*_*e*_ = 1.32 at the time of highest seasonal transmission in the NH (given by the phase of seasonality *ϕ*). None of the 595 filtered samples has *R*_*e*_ below one at peak seasonality during the 2026-27 NH flu season (Fig. S8): the median is 1.21; the interquartile range, 1.14-1.33; the 95% range, 1.06-1.82.

We also evaluated *R*_*e*_ at four hypothetical reintroduction dates over the 2026-27 season (Fig. 4). Most samples have *R*_*e*_ *>* 1 if reintroductions occur on January 1st (Fig. 4B) or April 1st (Fig. 4C). If instead reintroductions occur very early (October 1st, Fig. 4A) or very late (July 1st, Fig. 4D) in the influenza season, a substantial proportion of the samples have *R*_*e*_ *<* 1. For the sample that best fits the NH data, *R*_*e*_ is above one (purple circles in Fig. 4) at all four hypothetical reintroduction dates (from earliest to latest date, *R*_*e*_ = 1.18, 1.32, 1.15, 1.04). The second-best sample has *R*_*e*_ *>* 1. However, the third- and fourth-best samples have *R*_*e*_ *<* 1 for reintroductions on July 1st, and the fourth-best also on April 1st (Fig. 4). Fig. 4 also lists the probability that these reintroductions would generate an outbreak of at least 100 infections, calculated from 105 stochastic simulations of Yamagata transmission dynamics, for each of the four best-fitting samples. These probabilities would likely be lower had we included individual heterogeneity (33).

**Fig. 4.**
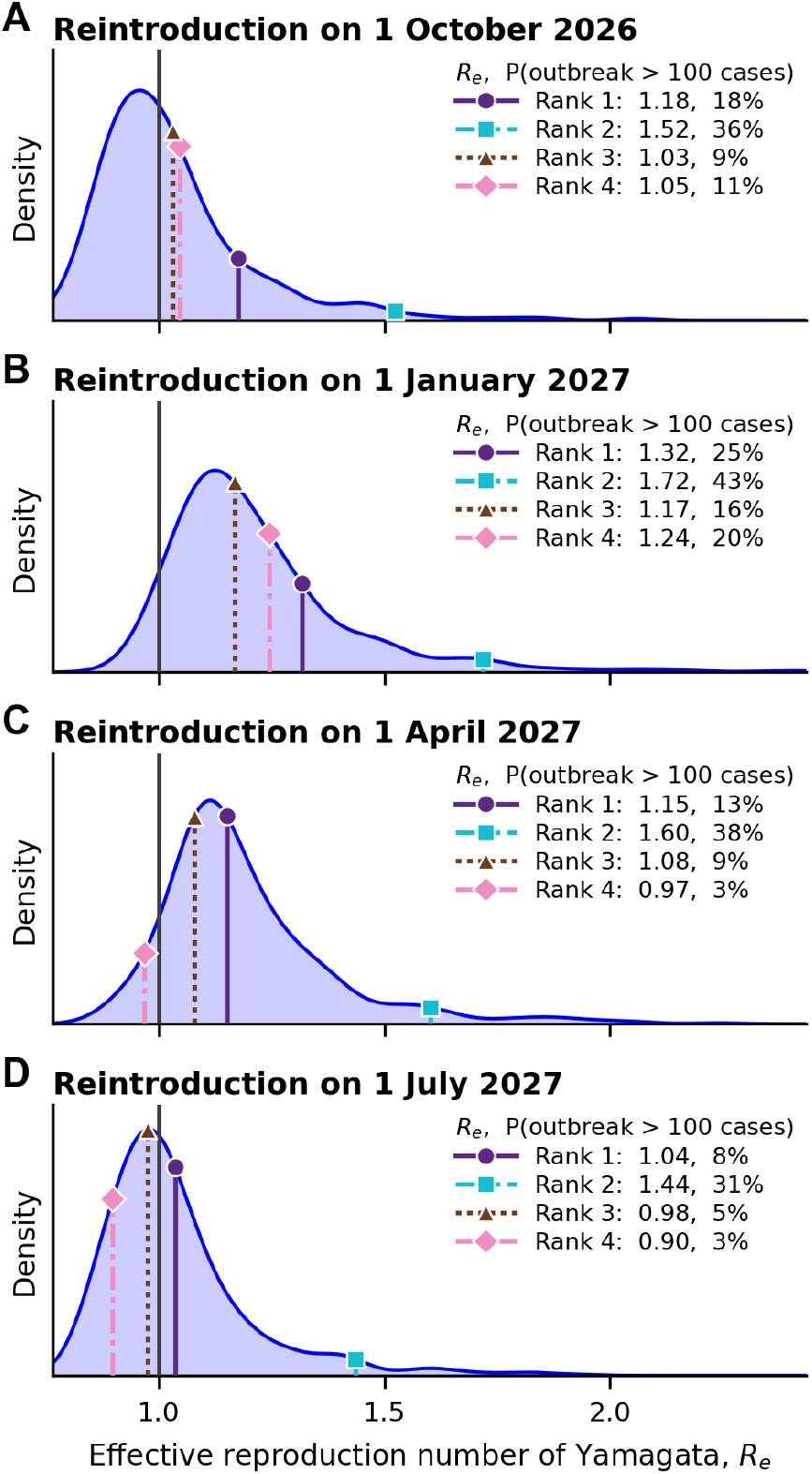
Distribution among the 595 filtered samples of *R*_*e*_, the effective reproduction number of Yamagata in the Northern Hemisphere region, at four hypothetical reintroduction dates over the 2026-27 influenza season. Lines and markers show these *R*_*e*_s for the four filtered samples that best fit the NH reported data (rank 1 is the best fit; Table S1), with, for each, the probability that a single reintroduction causes an outbreak of more than 100 cases (10^5^ stochastic simulations).

## Discussion

The mechanisms behind the disappearance of influenza B/Yamagata are currently unknown. This event represents a qualitative shift in the ecological dynamics of influenza B viruses. Given the large impact on human health caused by influenza B viruses, it is important to understand the potential implications of this extinction event. Thus, here we have examined the relative importance of epidemiological and immunological parameters in recapitulating the ecological patterns of influenza B observed in the last decade (2015-2025). We find that the likely cause of Yamagata extinction during the COVID-19 pandemic was the reduction of contacts during lockdowns coupled with its relatively low transmissibility (lower basic reproduction number than Victoria). We find little evidence to support other, primarily immunological or evolutionary, explanations. These findings suggest the pandemic pushed both lineages to very low prevalence, and Yamagata, the lineage with the lower intrinsic transmissibility, was unable to recover. This phenotypic difference between both lineages would be consistent with analyses which estimate a lower *effective* reproduction number for Yamagata than Victoria (20).

### Limitations

Despite the complexity of our mechanistic model, it has some important limitations. The model does not capture age structure, which can be important in shaping global persistence patterns (7). The age distribution of Yamagata cases was known to be bimodal, with higher incidence among adults than Victoria (which has a unimodal age distribution) (4). Thus, a higher adherence to social distancing measures in adults than children (34) could have contributed to the extinction of Yamagata (while allowing the persistence of Victoria).

Our three-region metapopulation structure (Northern Hemisphere, Tropics, and Southern Hemisphere) simplifies the real spatial connectivity of global influenza transmission (7, 8). Within each region, we assumed homogeneous mixing. Moreover, we also assumed that all three regions share the same intra-region contact rate and that interregion contact is governed by a single relative rate *m*. These assumptions may not capture the role of specific travel corridors, major transportation hubs, or within-region spatial heterogeneity in seeding and sustaining epidemics (4). More detailed metapopulation models with finer geographic resolution (35) could refine our estimates, particularly for the critical lockdown period when both lineages were at very low prevalence.

Our model did not include the impact of vaccines, which differ in efficacy each season (36, 37) and may elicit different immune responses depending on host immune history due to imprinting (30, 38, 39). Nonetheless, the vaccine efficacy was not substantially different between Yamagata and Victoria (36, 37), so we submit that vaccines were unlikely to have played a major role in the extinction of Yamagata. In the SI, we show that including vaccination at a constant *per capita* rate adjusts the basic reproduction numbers of both lineages by a multiplicative factor. This factor depends on the vaccine efficacy and coverage. The vaccine coverage was effectively the same between both lineages, as prior to the apparent extinction of Yamagata, WHO recommended quadrivalent influenza vaccines, with both Yamagata and Victoria strains (37). Thus, the basic reproduction numbers of both lineages would be adjusted by a similar factor, so the inclusion of vaccination should not change our main, qualitative conclusions. In fact, accounting for the loss of vaccine-derived immunity to Yamagata raises the estimates for its effective reproduction number over the 2026-27 influenza season (see SI).

We did not directly model pathogen evolution or reassortment (9). The parameters for each lineage are constant in time, and so our model does not directly capture phenotypic changes that the lineages might have undergone since 2015 (4). Antigenic drift (leading to evasion of host immunity) is one of the key features of global influenza epidemiology (7, 9). As a proxy for this antigenic evolution, we include the waning of immunity. However, this waning of immunity occurs at a constant rate (as in (26, 40)), and is independent of factors that may modulate the strength of antigenic drift each season (such as epidemic size (41) or antigenic cluster transitions (42, 43)).

Due to the noisiness of the data and the large, ten-dimensional parameter space, parameter estimation by maximum likelihood (31, 44) was not possible. Thus, instead, we evaluated a feature-based function identifying parameter combinations that were biologically plausible and recreated the qualitative patterns of influenza B lineages. This approach is similar to the methods adopted in (24) (latin hypercube sampling) and (45, 46) (feature-based selection of parameters). To quantify the importance of each parameter in generating the required ecological patterns, we used conditional SHAP values (47, 48). We also did not run stochastic simulations of the full epidemic dynamics, which might better recreate the patterns observed in the data.

### Comparison of model predictions to observed data

The large Yamagata epidemic in the 2017-18 winter season is reflected in the model output for the best-fitting parameter combination. In particular, the model predicts a relatively large Yamagata epidemic in that Northern Hemisphere influenza season, followed by smaller epidemics in the 2018-19 and 2019-20 seasons. These features qualitatively match the reported data (Fig. 1). Nevertheless, the predicted 2019-20 Yamagata epidemic is still notably larger than observed (Fig. S20). This discrepancy between the model and the data might be explained by the documented anomalous influenza dynamics of the 2019-20 season (26).

The third-best fitting sample (Fig. S15) mimics this observed pattern better, with a large outbreak in the 2017-18 season dominated by Yamagata. The model predicts very few Yamagata reports for the next two seasons, even fewer than in the observed data (Fig. S22). The Yamagata prevalence in the 2018-19 and 2019-20 seasons is smaller than it was in the 2015-16 and 2016-17 seasons, because many individuals had become infected with (and, subsequently, immune to) Yamagata during the 2017-18 outbreak. This high population immunity to Yamagata might have made it very vulnerable to extinction during COVID-19 (18). Running the deterministic simulation for this sample but without the lockdown confirms that Yamagata would not have gone extinct without the impact of the pandemic (Fig. S19). Nonetheless, Yamagata prevalence in this sample would have remained very low until the 2021-22 season, thus supporting the conclusion of (18) about the potential contribution of the 2017-18 season to Yamagata’s extinction. Unlike (18), we did not directly measure antigenic evolution. However, as seen in these samples, it is possible to recreate the observed qualitative patterns of influenza B without slower antigenic evolution in Yamagata. Instead, here these patterns appear due to the higher intrinsic transmissibility of Victoria.

### The potential for eradication of all influenza B viruses

The extinction of Yamagata increases population susceptibility to Victoria, assuming that Yamagata provided some partial cross-immunity to Victoria (*σ*_*Y V*_ *>* 0). As expected, after adjusting for the transient effects of the lockdown, in our model the absence of Yamagata increases the prevalence of Victoria (Table S1). The magnitude of this increase substantially depends on the strength of the cross-immunity from Yamagata to Victoria, which (19) suggests to be small. Nevertheless, for the four best-fitting samples, eliminating Yamagata substantially reduces the overall prevalence of influenza B (Table S1).

Given that Victoria was likely very close to extinction, it is interesting to consider the feasibility of its eradication. Using the estimates of *c*_0_ (the initial reduction in contacts) and *τ* (the lockdown timescale), we can obtain approximate benchmarks for a potential vaccination campaign. Neglecting the dynamics of immunity and population demography, the effect of COVID-19 control measures on the Victoria force of infection was a multiplicative factor 1 − *c*_0_*e*^*−T/τ*^, where *T* is the time since the implementation of lockdown. We can consider vaccination of a fraction *f* of the population, with a vaccine that initially reduces susceptibility by a factor *θ*_*S*_ ∈ [0, 1] and infectiousness by a factor *θ*_*I*_ ∈ [0, 1]. Thus, 1 − *θ*_*S*_*θ*_*I*_ is the transmission-blocking impact of the vaccine (49). Hence, the immunization campaign reduces the force of infection by a factor of 1 − *f* (1 − *θ*_*S*_*θ*_*I*_), assuming a pulse of vaccination (50). If vaccine immunity decays exponentially over time (51) with characteristic timescale *ν*, the aforementioned factor becomes 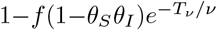 where *T*_*ν*_ is the time since vaccination. Thus, the effect of *f* (1 − *θ*_*S*_*θ*_*I*_) is analogous to the reduction in contacts *c*_0_, while the effect of *ν* is analogous to the lockdown timescale *τ*. Based on the best-fitting sample, Victoria was close to extinction with *c*_0_ = 0.34 and *τ* = 390 days. Hence, for a vaccine with characteristic duration of slightly over one year (*ν* ≈ 390 days) and with 1 − *θ*_*S*_*θ*_*I*_ ≈ 50% efficacy, vaccinating *f* ≈ *c*_0_*/*(1 − *θ*_*S*_*θ*_*I*_) = 67% of the global population could bring Victoria close to extinction. This tentative target, which should not be taken literally, would be achievable in high-income countries (37), but would represent a major challenge in the rest of the world.

Given the absence of any zoonotic spillover for Yamagata, laboratory escape represents the greatest potential risk for its reintroduction (52). The extinction of Yama-gata should not be interpreted as evidence that the human population is now protected against its hypothetical return. On the contrary, since 2020 its absence has allowed immunity to wane, while demographic turnover has increased the proportion of individuals with no prior exposure. Moreover, Yamagata has been removed from WHO’s vaccine recommendations (53). Some immunity to Yamagata is still being generated through cross-immunity from Victoria infections. However, we have found that this cross-immunity is not enough to compensate for the aforementioned decrease in population immunity to Yamagata. Our model predicts that, if reintroduced into human hosts, Yamagata would be likely to sustain transmission and cause a substantial epidemic, possibly returning to its prepandemic coexistence with Victoria. To reduce the probability of these undesirable outcomes, it would be prudent to handle Yamagata samples under biosafety level 3 (BSL3) conditions, rather than the BSL2 that is currently required (54).

## Materials and Methods

### Two-lineage single-region deterministic model

We used a deterministic compartmental model with homogeneous mixing. The epidemiological structure mimics an SIRS (32) model, with waning of immunity (representing antigenic drift) and population dynamics (with per capita birth rate *µ*, equal to the per capita death rate so the population is in demographic equilibrium). To capture the epidemic dynamics of both lineages (Victoria and Yamagata), we used a status-based formulation for immunity (55): *S*_*V*_ and *I*_*V*_ are the hosts susceptible to and infectious with Victoria. Using *N* for the population size, *N* −*S*_*V*_ −*I*_*V*_ hosts are immune to Victoria. Analogous compartments *S*_*Y*_, *I*_*Y*_ are defined for Yamagata. We further assumed that cross-immunity is polarized/all-or-none (56): this assumption is discussed in the SI. We let *σ*_*V Y*_ ∈ [0, 1] denote the probability that a host becomes immune to Yamagata upon recovery from Victoria (*σ*_*Y V*_ is defined analogously). Hence, the ODEs are:

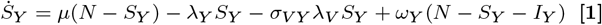

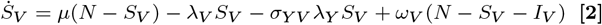

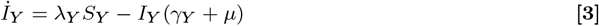

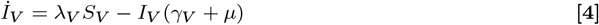

where, for * = *Y, V* denoting lineage, *λ*_*\**_ = (*β*_*\**_*/N*)*I*_*\**_ is the force of infection, *β*_*\**_ is the transmissibility, *γ*_*\**_ is the recovery rate, and *ω*_*\**_ is the waning rate (so *T*_*\**_ = 1*/ω*_*\**_ is the average duration of immunity to *). Equations 1-4 have a unique coexistence equilibrium, which is stable if it is biologically feasible (57). The basic reproduction number of the lineage * is *R*_0,*\**_ = *β*_*\**_*/*(*γ*_*\**_ + *µ*). We assumed both lineages have the same average duration of infection (1*/γ*_*\**_ = 3.4 days (58) as in (18)) and also fixed the average life expectancy at 1*/µ* = 75 years.

### Three-region global structure with seasonality

Following (7), we considered three global regions: temperate and sub-tropical countries of the Northern Hemisphere (population *N* ^*i*^ = 4 billion), Tropics (population *N* ^*ii*^ = 3.4 billion) and temperate and sub-tropical countries of the Southern Hemisphere (population *N* ^*iii*^ = 0.8 billion). We used Equations 1-4 for each region, adding global travel (see below) and seasonality outside of the tropics (modulated by a cosine function). Thus, in the Northern Hemisphere (NH) region, the transmission rate of lineage * ∈ {*Y, V*} depends on time *t* (given in days): 365.25 days

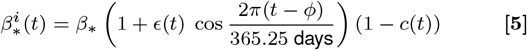

where *ϕ* is the phase of seasonality. The amplitude of seasonality *ϵ*(*t*) and the multiplier (1 − *c*(*t*)) are constants equal to *ϵ* and 1 before the COVID-19 lockdown. Their value after the emergence of COVID-19 is explained in the next subsection. Relative to the NH, seasonality is off phase in the Southern Hemisphere (SH) region. We assumed contacts between individuals of different regions occur at a relative rate *m*, so that, for example, the force of infection of lineage * in the Northern Hemisphere is

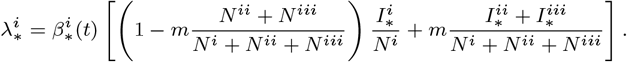

This expressions accounts for inter-region travel of both susceptible and infected hosts, ensuring that the (time-dependent) basic reproduction number in each region *i* remains 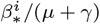 for both lineages * = *V, Y*.

#### Gradual relaxation of lockdown and return of seasonality

To capture the effects on influenza transmission of the COVID-19 lockdowns, we implemented an instantaneous reduction in transmission *c*_0_ and a gradual return to pre-pandemic transmission as in (10). Denoting the reduction in transmission by *c*(*t*) (acting on the force of infection as described above), we set *c*(*t*) = 0 for all *t < t*_*l*_ (where *t*_*l*_ is the time of the start of the lockdown, set to the end of March 2020). For *t* ≥ *t*_*l*_, we used *c*(*t*) = *c*_0_ exp (−(*t* − *t*_*l*_)*/τ*) where *τ* is the characteristic timescale of the lockdown. We also assumed that seasonality disappears at the start of lockdown and returns gradually (to account for the reduction in indoor activity), so that for *t > t*_*l*_ the amplitude of seasonality is *ϵ*(*t*) = *ϵ*(1 − exp (−(*t* − *t*_*l*_)*/τ*)). We fixed the baseline amplitude at *ϵ* = 0.1, so epidemic trajectories converge to a periodic attractor (59).

#### Latin Hypercube Sampling and feature-based search of samples

We allowed ten parameters of the model to vary within pre-specified upper and lower bounds (Table 1). We used a Latin Hypercube Sampling algorithm (27) to generate one million samples across the ten-dimensional space of variable parameters. First, we found the samples that allow for pre-pandemic coexistence. If a sample does not have a coexistence equilibrium according to single-region model (described above), we assume that coexistence is impossible in the global model (and hence we did not solve the ODEs). This eliminated a very small percentage of samples in which Yamagata and Victoria coexisted globally (due to the metapopulation structure of the three regions), but with at least one lineage not recreating the seasonal outbreaks observed in the data. If the sample does have a coexistence equilibrium in the single-region model, it can be proven to be unique (57) and we used its values as initial conditions (at time zero, assigned to January 1st, 2020) for each of the three global regions. We ran the deterministic model for *t*_*l*_ =20.25 years (i.e., until the COVID-19 lockdowns at the end of March 2020). We checked if either lineage had gone extinct before *t*_*l*_, defining a lineage * as extinct if it had less than one active infection in each of the three global regions (i.e., 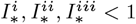). To allow for transient effects while the system reaches its periodic attractor (seasonality does not allow for a stationary equilibrium), when evaluating pre-pandemic coexistence we only declared a lineage extinct if the extinction condition was satisfied in the two years before the lockdown. If the condition for pre-pandemic coexistence was satisfied, we continued running the simulation, with the effects of COVID-19 as described in the previous subsection. If a lineage satisfied the extinction condition after March 2020, we set its prevalence to zero in all three regions. We defined the reemergence time of Victoria as the first week after its post-lockdown minimum in which the number of new Victoria infections in the NH region was equal to 10% of the average peak weekly number of new Victoria infections in the NH over the five influenza seasons before the pandemic (2015-16 to 2019-20). In the NH data, this remeergence threshold was met on Oct 18th, 2021. We selected the samples in which Victoria reemerged in the 2021-22 season (between the start of October 2021 and the end of March 2022), and Yamagata went extinct after March 2020 but before the reemergence of Victoria (as reflected in the data).

#### XGBoost and SHAP values

To quantify the contribution of each variable parameter (Table 1) to the selection outcome, we framed the problem as binary classification: each of the one million LHS samples was labeled positive (selected) or negative (not selected). The dataset is highly imbalanced, with only 3,844 positive samples (0.38%). To address this, we randomly downsampled the negative class to a 10:1 negative-to-positive ratio. We trained an XGBoost gradient-boosted tree classifier (28) using XGBClassifier from the xgboost package for Python. To interpret the trained classifier, we applied SHAP (SHapley Additive exPlanations) (29), using the TreeExplainer implementation from the shap package. SHAP decomposes each prediction into additive contributions from each feature based on Shapley values from cooperative game theory, accounting for feature interactions. We extracted SHAP values for the positive class (i.e., the probability of a sample being selected) across all training samples. For each parameter, we report the mean absolute SHAP value as a summary measure of overall feature importance (Table 1); a beeswarm plot (Fig. S3) additionally shows the direction and spread of each parameter’s influence.

#### Likelihood function

We constructed a likelihood function (31) to compare, for each sample, the deterministic output against the reported data in the NH region. The number of infections reported each week *w* is denoted with *O*_*V*_ (*w*) for Victoria infections, *O*_*Y*_ for Yamagata infections, and *O*_*U*_ for untyped infections (unspecified lineage). We defined a weekly typing probability, *p*_*w*_, equal to the number of typed infections, *O*_*T*_ = *O*_*V*_ + *O*_*Y*_, divided by all reported infections in a given week *w, O*_*A*_ = *O*_*T*_ + *O*_*U*_. We assumed that the reporting rate *ρ*_*w*_ (equal for both lineages) is either *ρ*_*−*_ and *ρ*_+_, depending on whether the week *w* is before or after the week *w*_*†*_ of July 1st, 2020 (to represent, after an adjustment period, increased testing and reporting of influenza-like illnesses since COVID-19). Further, we assumed that weekly counts were subject to a Poisson-distributed reporting error (as in (26)). Thus, if for a sample *s* the deterministic model predicts *P*_*\**_(*w*; *s*) new infections of lineage * in the NH region during week *w*, the reported number is a Poisson-distributed random variable with mean *p*_*w*_ *ρ*_*w*_ *P*_*\**_(*w*; *s*). Similarly, the number of untyped reported infections in the NH is a Poisson-distributed random variable with mean (1 − *p*_*w*_)*ρ*_*w*_*P*_*A*_(*w*; *s*), where *P*_*A*_ = *P*_*V*_ + *P*_*Y*_ are all predicted infections.

Thus, the likelihood function for a sample *s* is

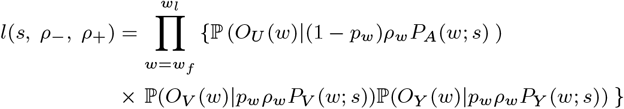

where *w*_*f*_, *w*_*l*_ are the first and last weeks of the time period considered (October 1st, 2015, to September 30th, 2025) and ℙ(·|Λ) is the Poisson probability mass function with parameter Λ. The Poisson distribution means that, for each sample *s*, the likelihood is maximized for the following reporting rates:

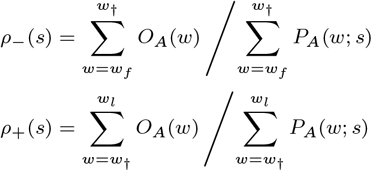

The distributions for the inferred reported rates over all 595 samples are shown on Fig S10 and S11. We evaluated each sample *s* with the likelihood *l*(*s, ρ*_*−*_(*s*), *ρ*_+_(*s*)).

#### Stochastic dynamics of Victoria reemergence

We initiated each stochastic realization with the (rounded) values of the deterministic variables 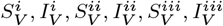 at the moment of Yamagata extinction. We then simulated transmission dynamics using the tau-leaping algorithm (32), with time step Δ*t* = 0.1 day. Within each time step, the number of events of each type in each region was drawn independently from a Poisson distribution. For example, the number of new infections in the NH region is given by a Poisson-distributed random variable with mean 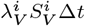. After sampling all events, the population states were updated according to the transmission dynamics described earlier. We truncated the number of realised events so that the model compartments never reached negative values. We declared Victoria extinct in a given realization when the total number of active infections across all three regions was zero at the reemergence time (as defined earlier based on the deterministic trajectory). Each realization ran until September 30th, 2026.

#### Stochastic dynamics of Yamagata reintroduction

The effective reproduction number of Yamagata in the Northern Hemisphere at a hypothetical reintroduction time is

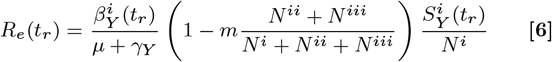

where 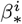 is given by Equation 5 and *S*^*i*^ is given by the deterministic model for Victoria dynamics. We calculated the probability that a reintroduction causes an outbreak of at least 100 infections by running 10^5^ independent stochastic simulations. We initialized each simulation with only one Yamagata infection in the NH region. The susceptible Yamagata compartment in each region was initialized with the rounded values from the deterministic model. Since each simulation was limited to 100 concurrent Yamagata infections, we assumed that the Yamagata outbreak does not affect Victoria dynamics, which we assumed followed the deterministic solution (Fig. 3). The Victoria force of infection determines the rates at which individuals become immune to Yamagata through cross-immunity: e.g., rate 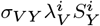 in the NH region. These rates were used to produce the stochastic trajectories, via Poisson tau-leaping (as earlier for the stochastic dynamics of Victoria).

#### Data, Materials, and Software Availability

We obtained the weekly influenza B data from WHO’s FluNet (Influenza Laboratory Surveillance Information) (21). We downloaded the data separately for the three possible values of the field ‘Tropics’: Northern Hemisphere Temperate and Sub-Tropical (generating FluNet NH.csv), Tropical (generating FluNet Tr.csv), and Southern Hemisphere Temperate and Sub-Tropical (generating FluNet SH.csv). These files along with our source code are available online in a github repository github.com/maria-a-gutierrez/Yamagata-extinction.

## Data Availability

All relevant data and code are available online in a Github repository

https://github.com/maria-a-gutierrez/Yamagata-extinction

## ACKNOWLEDGMENTS

We thank the Rohani Lab for comments. This project has been funded with Federal funds from the National Institute of Allergy and Infectious Diseases, National Institutes of Health, Department of Health and Human Services, under Contract No. 75N93021C00018 (NIAID Centers of Excellence for Influenza Research and Response, CEIRR).

## Supporting Information for

### Supporting Information Text

#### Vaccination

To understand if vaccination may change qualitatively our findings, here we extend the two-lineage single-region deterministic model to include vaccination. In particular, we add a term −*rθ*_*Y*_ *S*_*Y*_ to the expression for 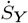 (Equation 1 of the main text), where *r* is the per capita rate of vaccination of individuals susceptibles to Victoria and *θ*_*Y*_ is the probability that a susceptible host becomes immune to Yamagata upon vaccination (as with cross-immunity, we assume that vaccine-induced immunity is also of the all-or-none type). We also assume that vaccine-induced immunity wanes at the same rate *ω*_*Y*_ = 1*/T*_*Y*_ as infection-induced immunity (so the term *ω*_*Y*_ (*N* − *S*_*Y*_ − *I*_*Y*_) in Eq. 1 remains unchanged). Finally, we do not allow vaccination of infectious individuals, so the equation for *İ*_*Y*_ also remains unchanged. At the disease-free equilibrium, the number of susceptible hosts to Yamagata is *S*_*Y*_ = *N* (*µ* + *ω*_*Y*_)*/*(*µ* + *ω*_*Y*_ + *rθ*_*Y*_). Thus, for *R*_0,*Y*_ = *β*_*Y*_ */*(*γ* + *µ*) as before, the initial effective reproduction number of Yamagata is

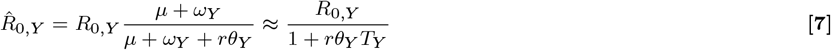

using *µ* ≪ *ω*_*Y*_ = 1*/T*_*Y*_ (since for the values considered in our analysis, the average duration of immunity is much shorter than the average host lifespan). Similarly, the initial effective reproduction number of Victoria is

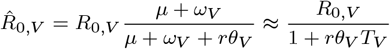

where *θ*_*V*_ is the probability that a susceptible host becomes immune to Victoria upon vaccination. We use the same per capita vaccination rate *r* because prior to the apparent extinction of Yamagata, influenza vaccines were quadrivalent, with both a Yamagata strain and a Victoria strain (37). If we apply the results of the main text to the initial effective reproduction numbers 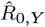 and 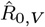 instead of *R*_0,*Y*_ and *R*_0,*V*_, we get that 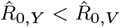 is important to recreate the ecological patterns of influenza B. Moreover, assuming the same both lineages had the same vaccine efficacy *θ* = *θ*_*V*_ = *θ*_*Y*_ (37), we get

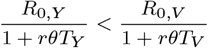

Among the ‘selected’ samples, we have *T*_*Y*_ ≈ *T*_*V*_ (see Table 1 of the main text), so we recover *R*_0,*Y*_ *< R*_0,*V*_. Moreover, in the ‘filtered’ samples we imposed *T*_*Y*_ *< T*_*V*_ based on evolutionary grounds (4), so even after accounting for vaccination we still have *R*_0,*Y*_ *< R*_0,*V*_ (higher intrinsic transmissibility for Victoria than Yamagata).

Equation 7 can also adjust the effective reproduction number of Yamagata during the 2026-27 NH influenza season to account for the removal of Yamagata strains from seasonal influenza vaccines. Since our model assumes that vaccine immunity is factored into the estimates of the reproduction number, the result of the main text means that

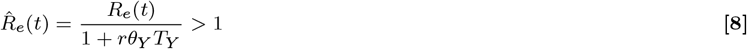

where 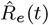 is the peak 2026-27 effective reproduction number of Yamagata including vaccination. Therefore, *R*_*e*_(*t*), the actual peak 2026-27 effective reproduction number of Yamagata (assuming all vaccine-derived immunity to Yamagata has waned), is still greater than one: *R*_*e*_(*t*) *>* 1 + *rθ*_*Y*_ *T*_*Y*_.

### All-or-none immunity assumption

The two-lineage status-based model that we use to evaluate the one million samples assumes all-or-none cross-immunity. This assumptions means that only four variables are tracked (for each region): *S*_*V*_, *S*_*Y*_, *I*_*V*_, and *I*_*Y*_, because all hosts in *S*_*\**_ are fully susceptible to lineage *. Changing this assumption (but keeping the status-based formulation) to instead consider leaky susceptibility requires an additional two compartments (for each region): *P*_*V*_ and *P*_*Y*_, tracking the hosts partially susceptible to lineage *V* and *Y* (appendix of (57)). We ran this otherwise equivalent model on the 595 filtered samples, finding that 187 of them no longer recreate the qualitative features of influenza B. It is highly likely that some of the samples rejected with the all-or-none formulation would be selected under leaky immunity. Of the 187 that are no longer selected, Yamagata never goes extinct in 116. This is consistent with leaky immunity generally permitting more transmission, as all hosts with leaky immunity will eventually be infected given enough exposure events. Nevertheless, we do not expect that leaky immunity would substantially change the qualitative patterns described in this manuscript. Among the samples that still match the qualitative features of influenza B, Victoria has a higher basic reproduction number than Yamagata in 99% of them (97% with all-or-none immunity) and the minimum Victoria prevalence changes by a median factor of 1.01. Moreover, the qualitative behavior of the four best-fitting samples remains the same with leaky immunity (Figs. S12, S14, S16, S18).

**Fig. S1.**
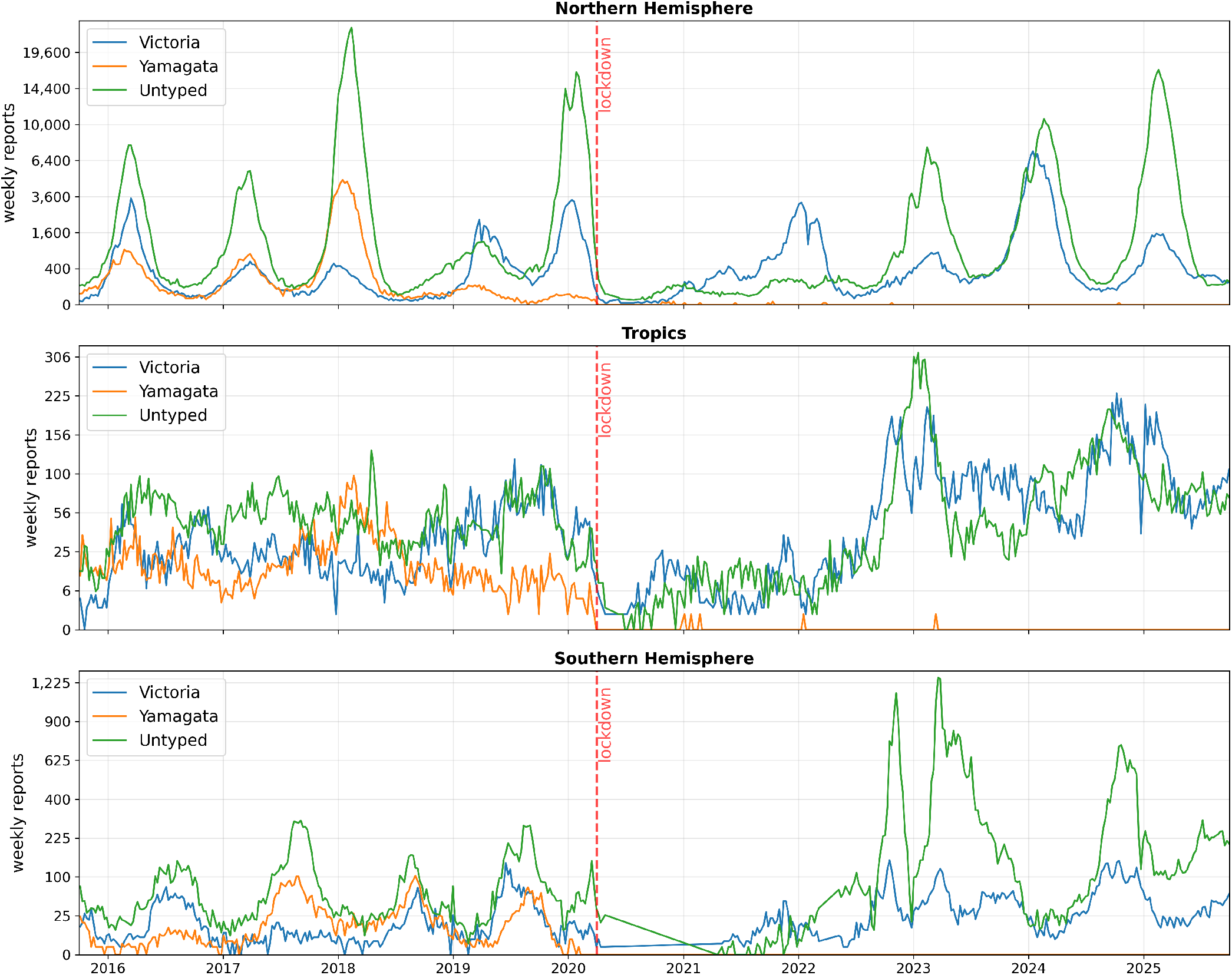
Global data from FluNet for influenza B: as Fig. 1 in the main text but including the Tropics and Southern Hemisphere regions.

**Fig. S2.**
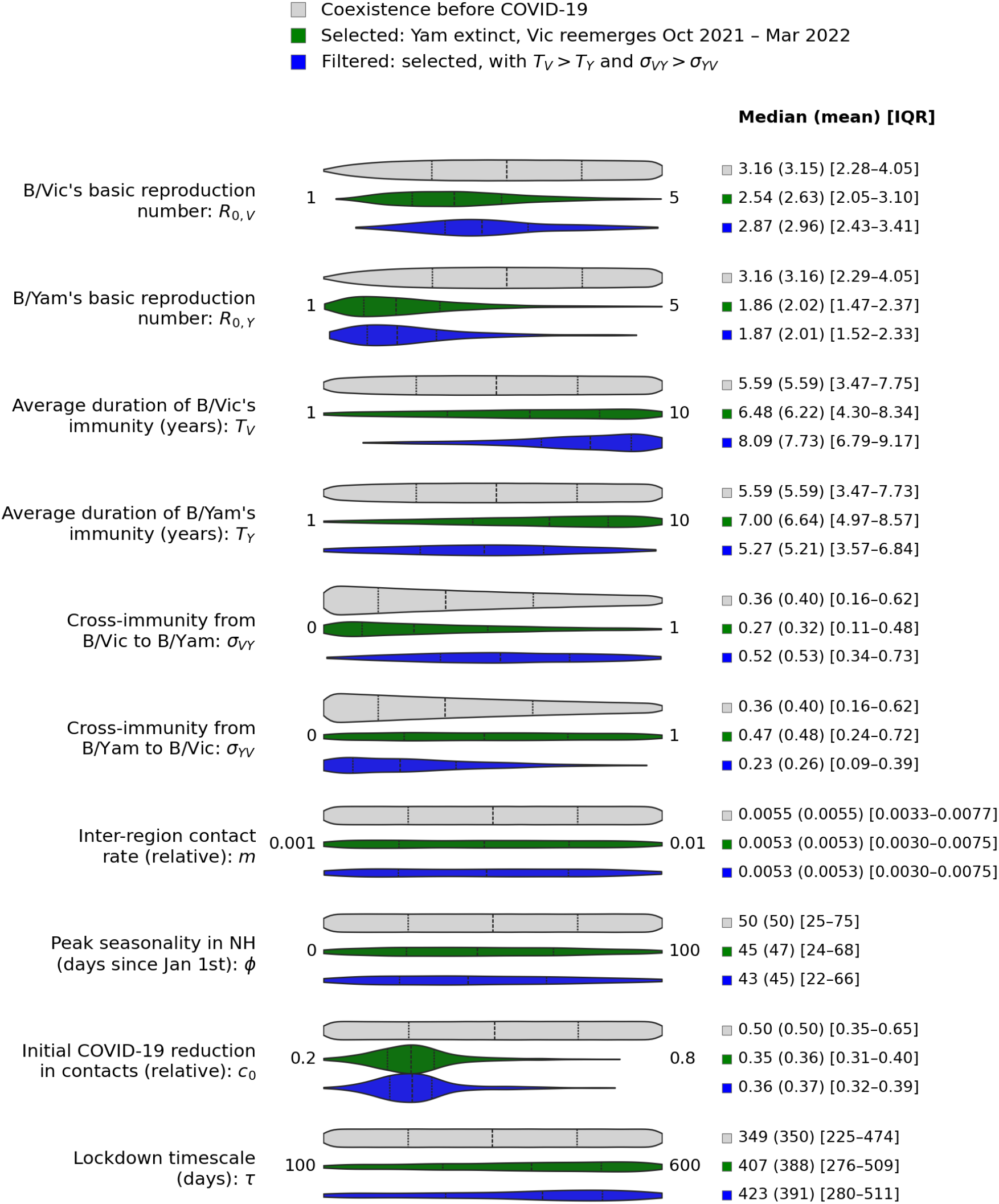
Parameter distributions for the ten variable parameters, among LHS-generated samples for which our model has (i, gray) Victoria and Yamagata coexisting prior to 2020, (ii, green) in addition to (i), Yamagata extinction prior to Victoria recovery (from Oct 2021 to March 2022), and (iii, blue) in addition to (ii), filtered biological features (*T*_*V*_ *> T*_*Y*_ and *σ*_*V Y*_ *> σ*_*Y V*_). The numbers at the left and right of the distributions are the respective lower and upper bounds used to generate the samples. Dashed lines on each violin plot mark the 25th, 50th (i.e., median), and 75th percentiles of the distribution. The column on the right gives, for each distribution, the median, the mean (in parentheses) and the interquartile range (in brackets).

**Fig. S3.**
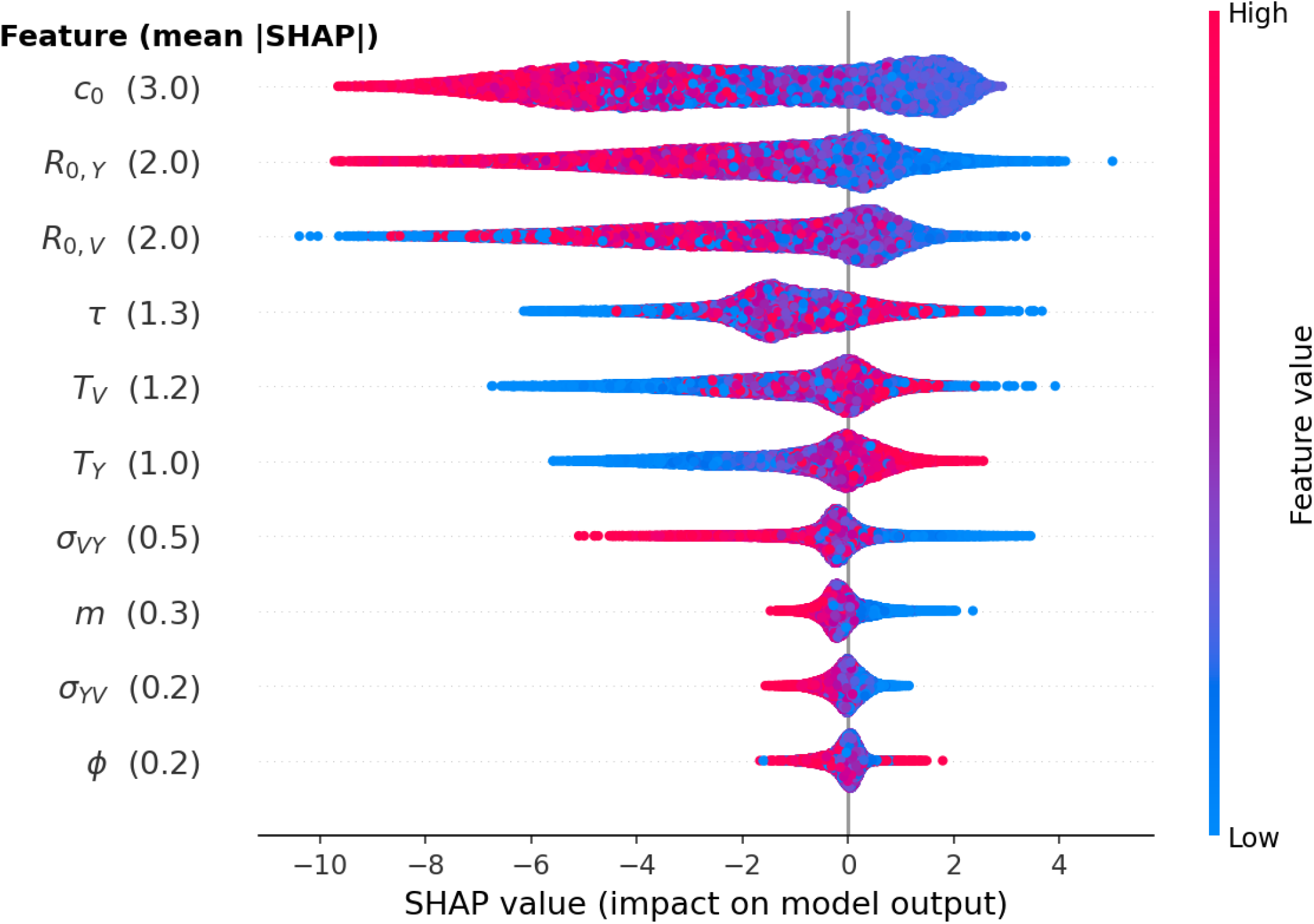
SHAP (SHapley Additive exPlanations) values for the XGBoost classifier (see Materials and Methods), quantifying how each parameter (‘feature’) influences the probability that a sample is selected (i.e., that it reproduces both the pre- and post-pandemic patterns of influenza B). Each point is one sample: its horizontal position is that parameter’s SHAP value for the sample (positive values push the prediction towards selection, negative away from it), and its color encodes the parameter value (blue: low, red: high). The parameters are displayed in descending order of their mean absolute SHAP value (shown in parentheses, and also listed in Table 1 of the main text), which is a summary measure of the parameter’s overall importance.

**Fig. S4.**
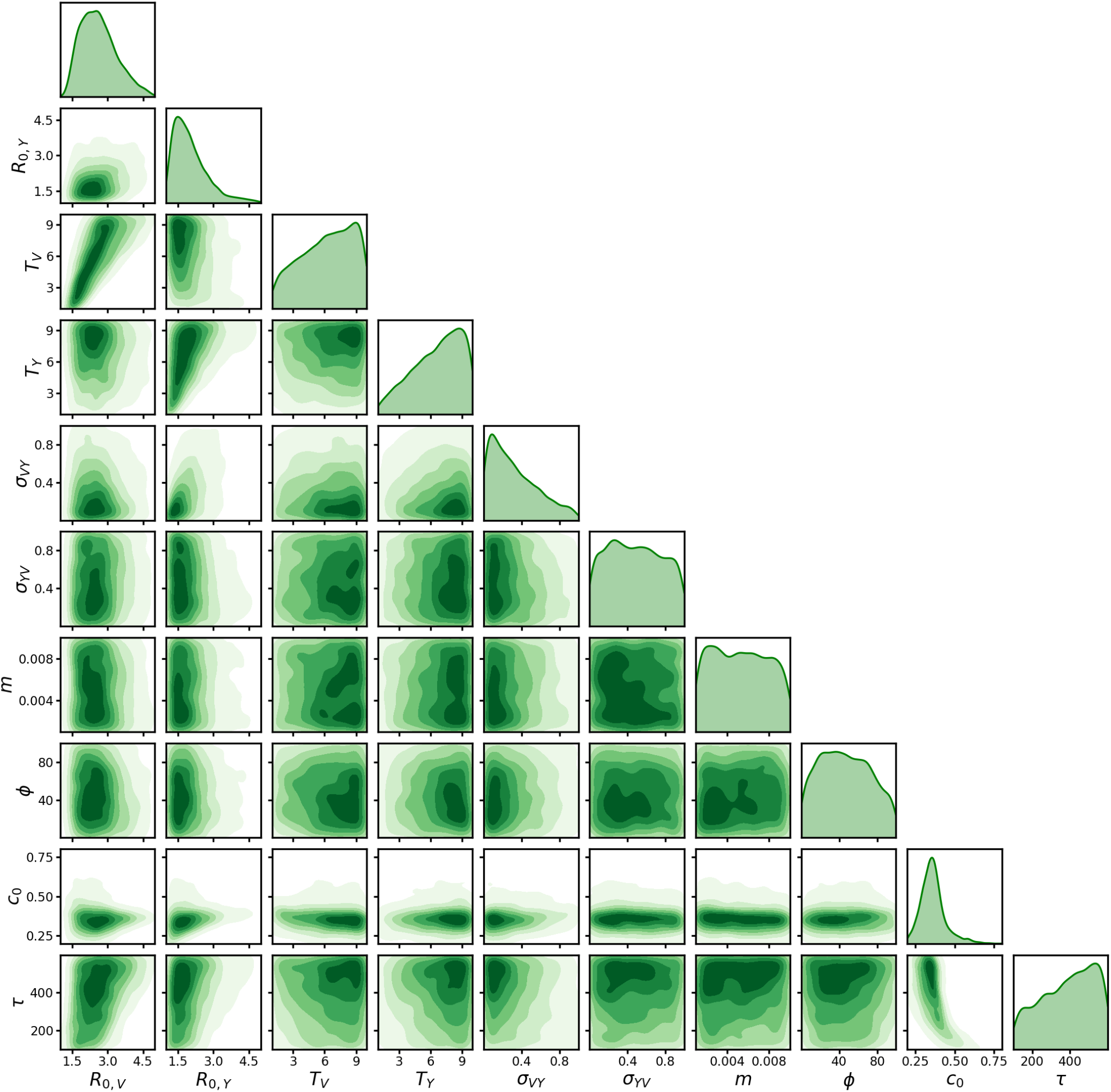
Pairwise distributions of the selected samples (green in Fig. S2). Panels below the diagonal show the bivariate kernel density of each pair of parameters, with the lowest 5% left blank. Panels on the diagonal show each parameter’s own density. Axes span the sampling bounds (Table 1 of the main text).

**Fig. S5.**
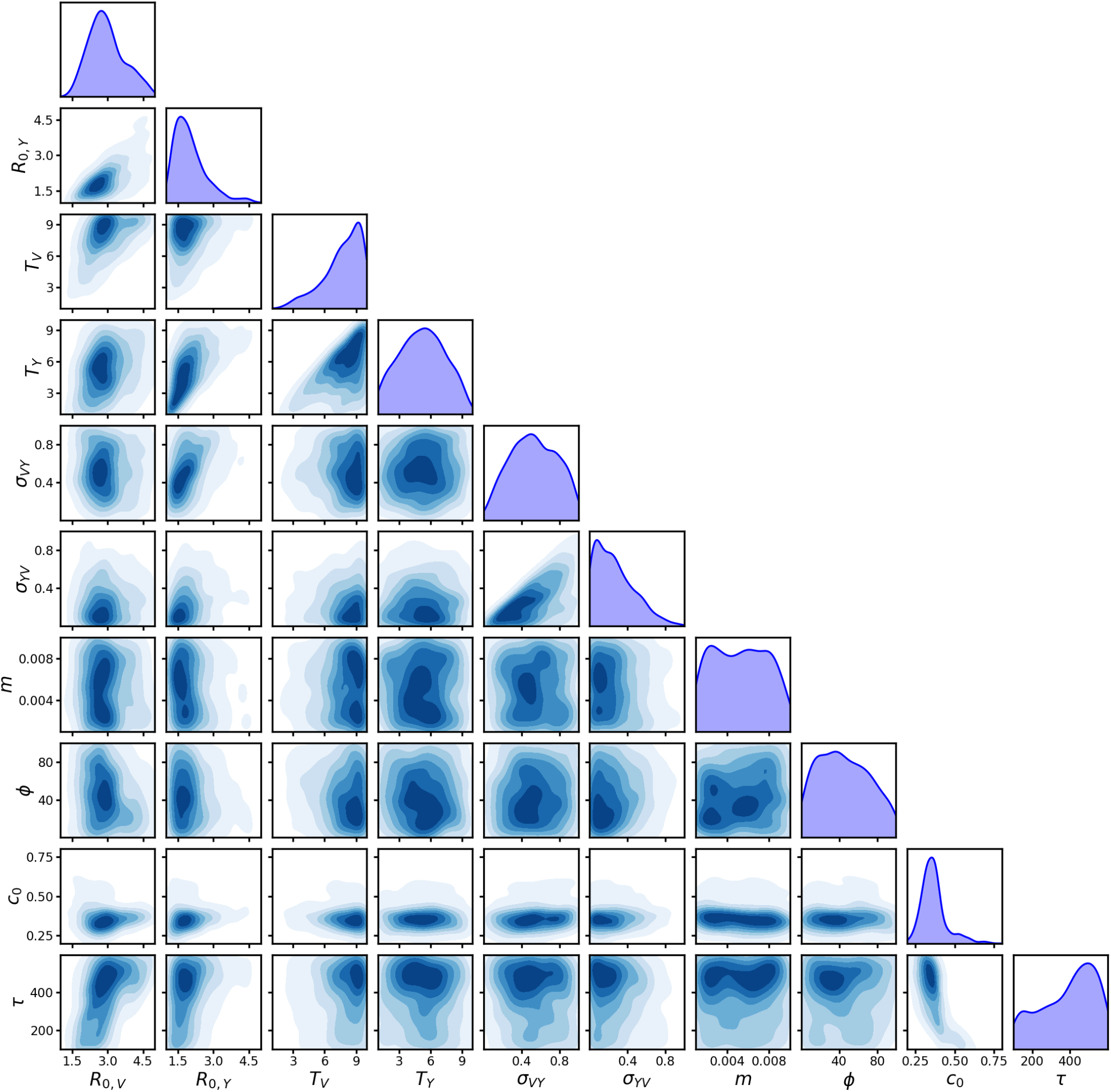
As Fig. S4, but showing only the biologically-filtered samples (blue in Fig. S2).

**Fig. S6.**
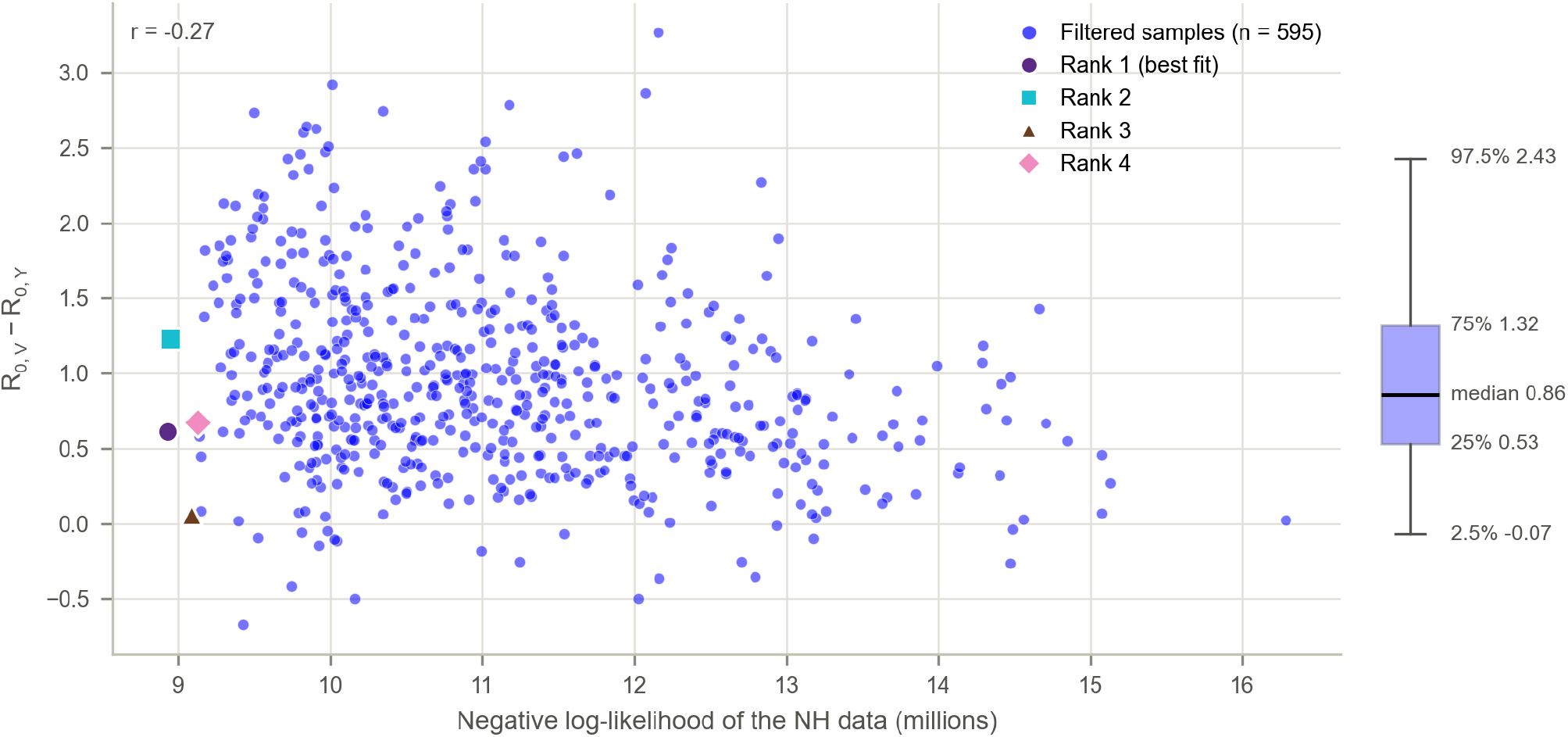
Difference *R*_0,*V*_ *− R*_0,*Y*_ for all 595 biologically-filtered samples, against their negative log-likelihood for the NH data (Spearman correlation coefficient r=-0.27). Markers show the four best fits to the NH data (Table S1). The box at the right marks the median, interquartile range and 95% range (2.5th to 97.5th percentiles) of the 595 samples. *R*_0,*V*_ *> R*_0,*Y*_ in 97% of the samples.

**Fig. S7.**
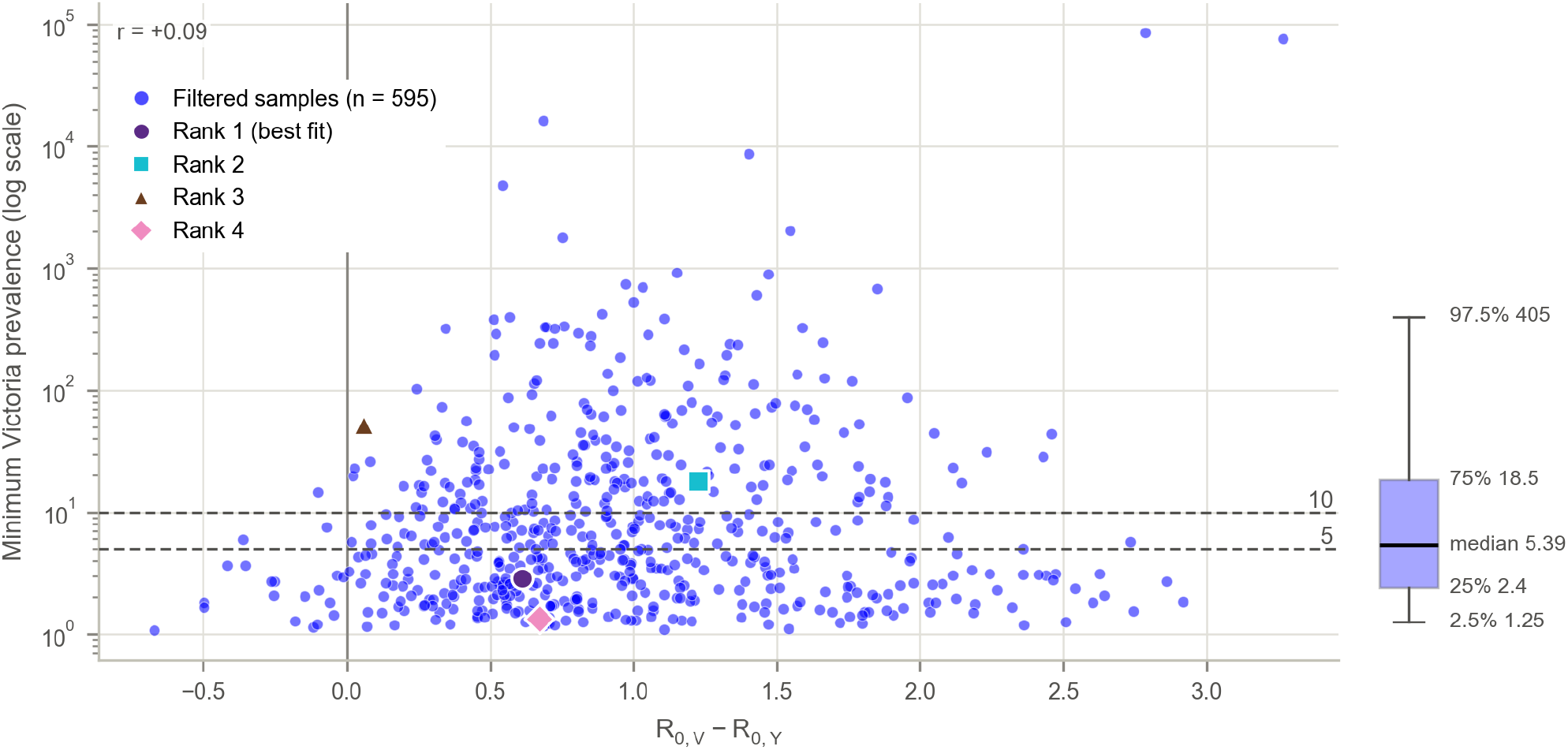
Minimum Victoria prevalence (deterministic) for all 595 biologically-filtered samples, against their difference *R*_0,*V*_ *− R*_0,*Y*_ (Spearman correlation coefficient r=0.09). The dashed lines mark 5 and 10 infections and the vertical line *R*_0,*V*_ = *R*_0,*Y*_. Markers show the four best fits to the NH data (Table S1). The box at the right marks the median, interquartile range and 95% range (2.5th to 97.5th percentiles) of the 595 samples.

**Fig. S8.**
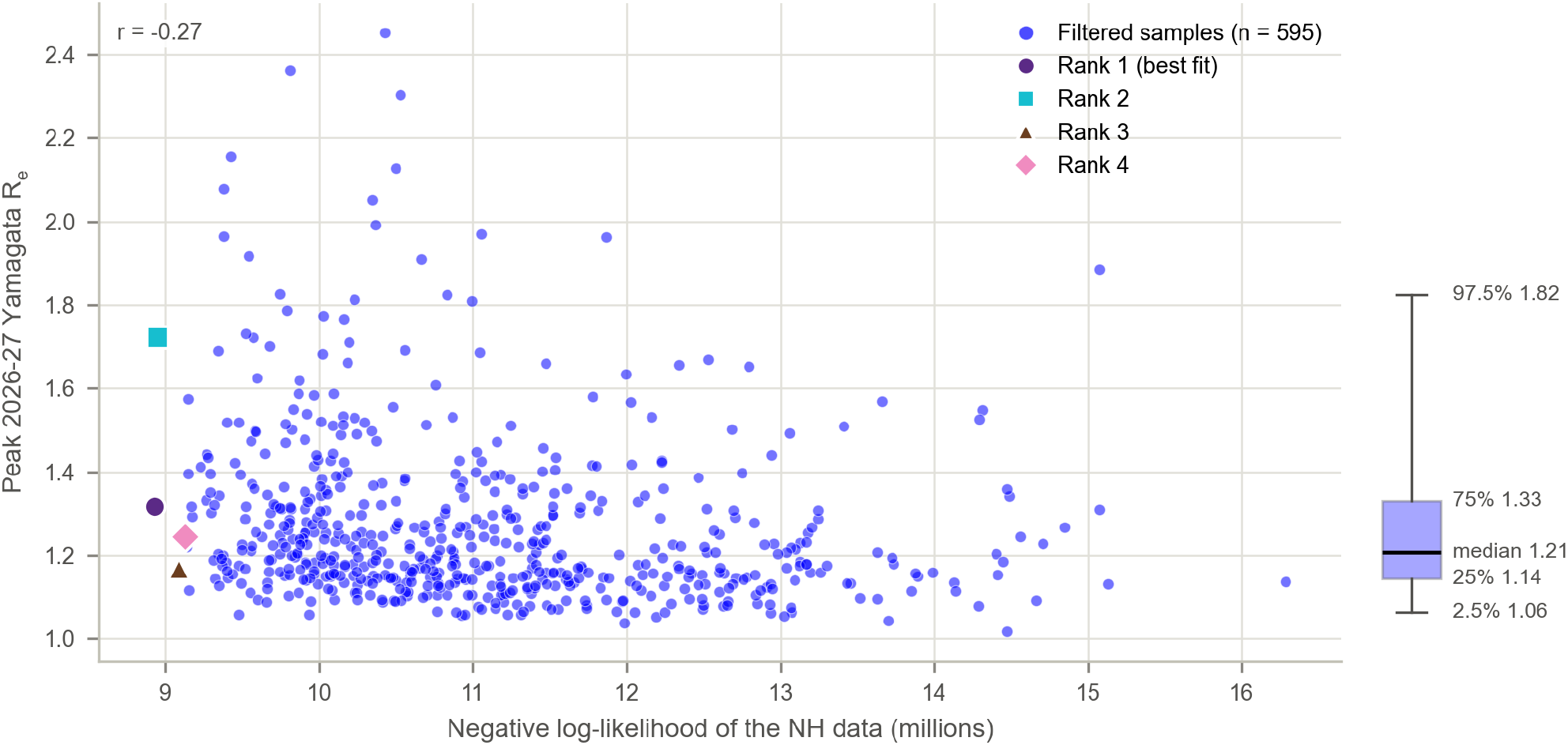
Effective reproduction number for Yamagata at the peak of the 2026-27 winter season for all 595 biologically-filtered samples, against their negative log-likelihood for the NH data (Spearman correlation coefficient r=-0.27). Markers show the four best fits to the NH data (Table S1). The box at the right marks the median, interquartile range and 95% range (2.5th to 97.5th percentiles) of the 595 samples.

**Fig. S9.**
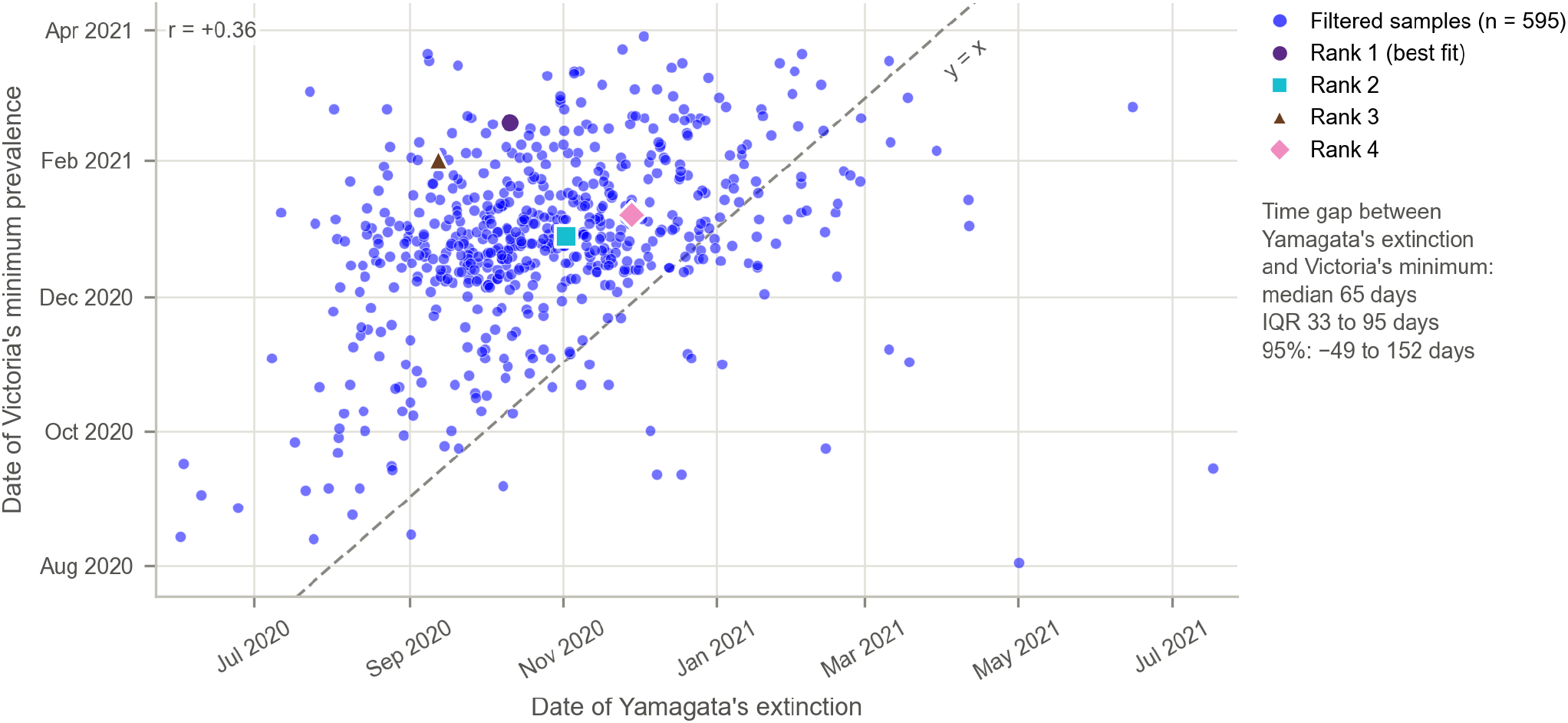
Date of the minimum global prevalence of Victoria (deterministic) for all 595 biologically-filtered samples against the date of Yamagata extinction (Spearman correlation coefficient r=0.36). Samples above the dashed line (*y* = *x*) have the minimum of Victoria after Yamagata extinction (Victoria’s minimum occurs first in only 56 of these 595 samples). Markers show the four best fits to the NH data (Table S1).

**Fig. S10.**
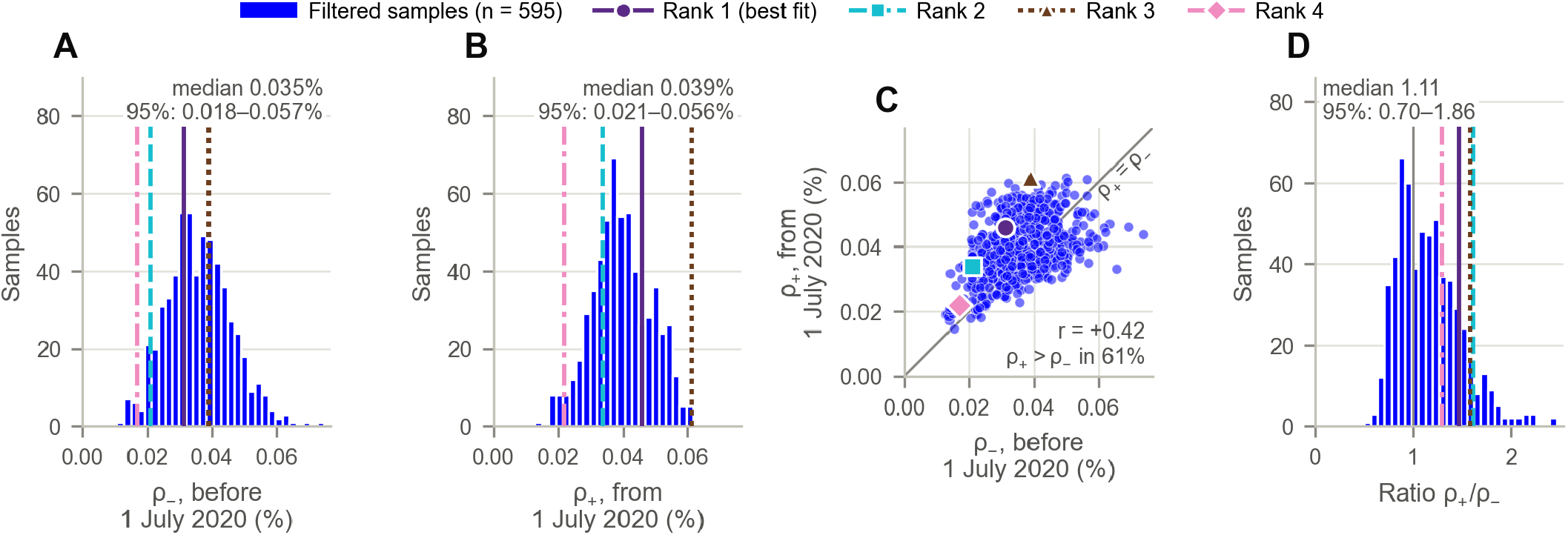
Reporting rates, *ρ*_*−*_ (until 30 June 2020) and *ρ*_+_ (from 1 July 2020) of the 595 biologically-filtered samples. (A, B) Distributions of *ρ*_*−*_ and *ρ*_+_, with their medians and 95% ranges. (C) *ρ*_+_ against *ρ*_*−*_ (Spearman correlation coefficient r=0.42), with the line *ρ*_+_ = *ρ*_*−*_ (gray). (D) Distribution of the ratio *ρ*_+_*/ρ*_*−*_, with the line *ρ*_+_ = *ρ*_*−*_ (gray). Lines and markers show the four best fits to the NH data (Table S1).

**Fig. S11.**
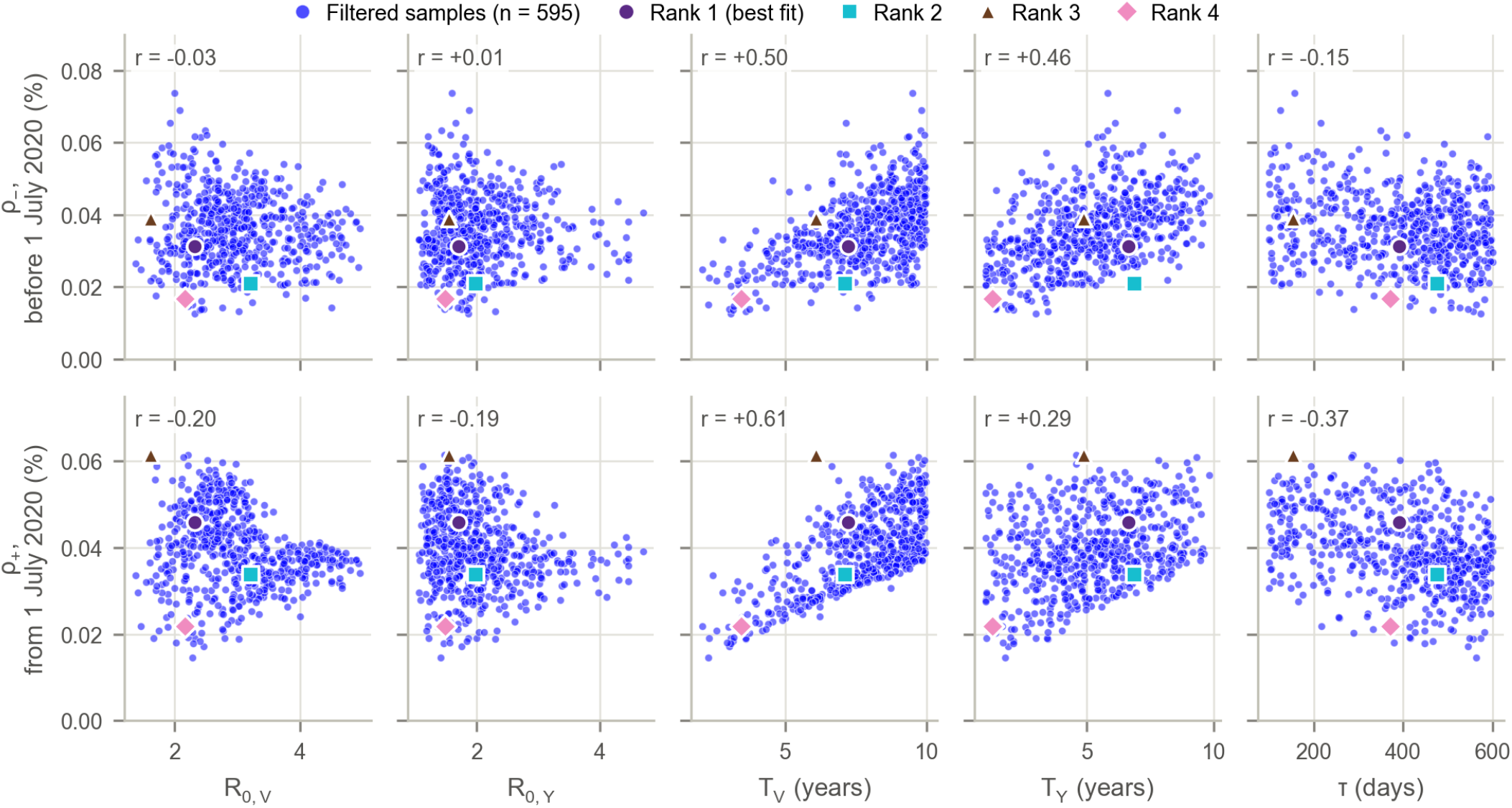
Reporting rates *ρ*_*−*_ (top row) and *ρ*_+_ (bottom row) of the 595 biologically-filtered samples against *R*_0,*V*_, *R*_0,*Y*_, *T*_*V*_, *T*_*Y*_ and *τ* (Spearman correlation coefficient r in each panel). Markers show the four best fits to the NH data (Table S1).

**Table S1.**
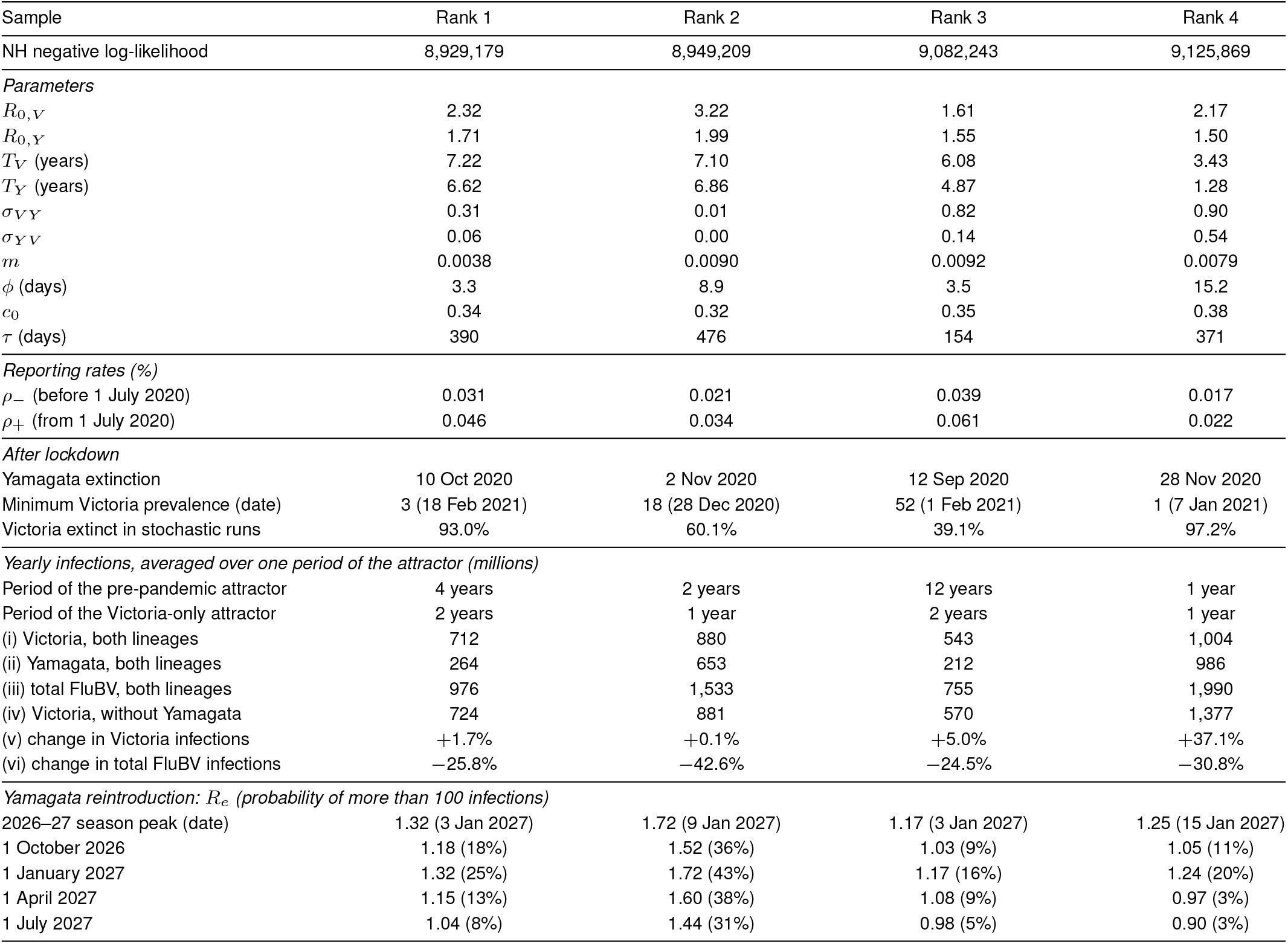
The four best fits to the NH data (lowest negative log-likelihood, NLL) among the 595 biologically-filtered samples. Infections (i)-(iv) are from the deterministic model, averaged over one period of the single- or two-lineage attractor, as appropriate. Infections with both lineages, (i)–(iii), are without the lockdown (contacts and seasonality as before 2020), from 1 October 2015 (15.75 years after the start of the simulation) so that the system has settled into the two-lineage periodic attractor. Victoria-only infections (iv) are from the model as fitted (with the lockdown, after which Yamagata dies out), from 1 January 2036 (15.75 years after the start of the lockdown), so that Victoria has settled into its single-lineage periodic attractor after the transient effects of the lockdown and of Yamagata’s extinction. The change (v) compares (iv) with (i): it is the effect of Yamagata’s absence on Victoria. The change (vi) compares (iv) with (iii): it is the net effect of Yamagata’s absence on influenza B (FluBV) infections.

| Sample | Rank 1 | Rank 2 | Rank 3 | Rank 4 |
|---|---|---|---|---|
| NH negative log-likelihood | 8,929,179 | 8,949,209 | 9,082,243 | 9,125,869 |
| *Parameters* |  |  |  |  |
| *R*<sub>0,*V*</sub> | 2.32 | 3.22 | 1.61 | 2.17 |
| *R*<sub>0,*Y*</sub> | 1.71 | 1.99 | 1.55 | 1.50 |
| *T*<sub>*V*</sub> (years) | 7.22 | 7.10 | 6.08 | 3.43 |
| *T*<sub>*Y*</sub> (years) | 6.62 | 6.86 | 4.87 | 1.28 |
| *σ*<sub>*VY*</sub> | 0.31 | 0.01 | 0.82 | 0.90 |
| *σ*<sub>*YV*</sub> | 0.06 | 0.00 | 0.14 | 0.54 |
| *m* | 0.0038 | 0.0090 | 0.0092 | 0.0079 |
| *ϕ* (days) | 3.3 | 8.9 | 3.5 | 15.2 |
| *c*<sub>0</sub> | 0.34 | 0.32 | 0.35 | 0.38 |
| *τ* (days) | 390 | 476 | 154 | 371 |
| *Reporting rates (%)* |  |  |  |  |
| *ρ*<sub>−</sub> (before 1 July 2020) | 0.031 | 0.021 | 0.039 | 0.017 |
| *ρ*<sub>+</sub> (from 1 July 2020) | 0.046 | 0.034 | 0.061 | 0.022 |
| *After lockdown* |  |  |  |  |
| Yamagata extinction | 10 Oct 2020 | 2 Nov 2020 | 12 Sep 2020 | 28 Nov 2020 |
| Minimum Victoria prevalence (date) | 3 (18 Feb 2021) | 18 (28 Dec 2020) | 52 (1 Feb 2021) | 1 (7 Jan 2021) |
| Victoria extinct in stochastic runs | 93.0% | 60.1% | 39.1% | 97.2% |
| *Yearly infections, averaged over one period of the attractor (millions)* |  |  |  |  |
| Period of the pre-pandemic attractor | 4 years | 2 years | 12 years | 1 year |
| Period of the Victoria-only attractor | 2 years | 1 year | 2 years | 1 year |
| (i) Victoria, both lineages | 712 | 880 | 543 | 1,004 |
| (ii) Yamagata, both lineages | 264 | 653 | 212 | 986 |
| (iii) total FluBV, both lineages | 976 | 1,533 | 755 | 1,990 |
| (iv) Victoria, without Yamagata | 724 | 881 | 570 | 1,377 |
| (v) change in Victoria infections | +1.7% | +0.1% | +5.0% | +37.1% |
| (vi) change in total FluBV infections | −25.8% | −42.6% | −24.5% | −30.8% |
| *Yamagata reintroduction: R<sub>e</sub> (probability of more than 100 infections)* |  |  |  |  |
| 2026–27 season peak (date) | 1.32 (3 Jan 2027) | 1.72 (9 Jan 2027) | 1.17 (3 Jan 2027) | 1.25 (15 Jan 2027) |
| 1 October 2026 | 1.18 (18%) | 1.52 (36%) | 1.03 (9%) | 1.05 (11%) |
| 1 January 2027 | 1.32 (25%) | 1.72 (43%) | 1.17 (16%) | 1.24 (20%) |
| 1 April 2027 | 1.15 (13%) | 1.60 (38%) | 1.08 (9%) | 0.97 (3%) |
| 1 July 2027 | 1.04 (8%) | 1.44 (31%) | 0.98 (5%) | 0.90 (3%) |

**Fig. S12.**
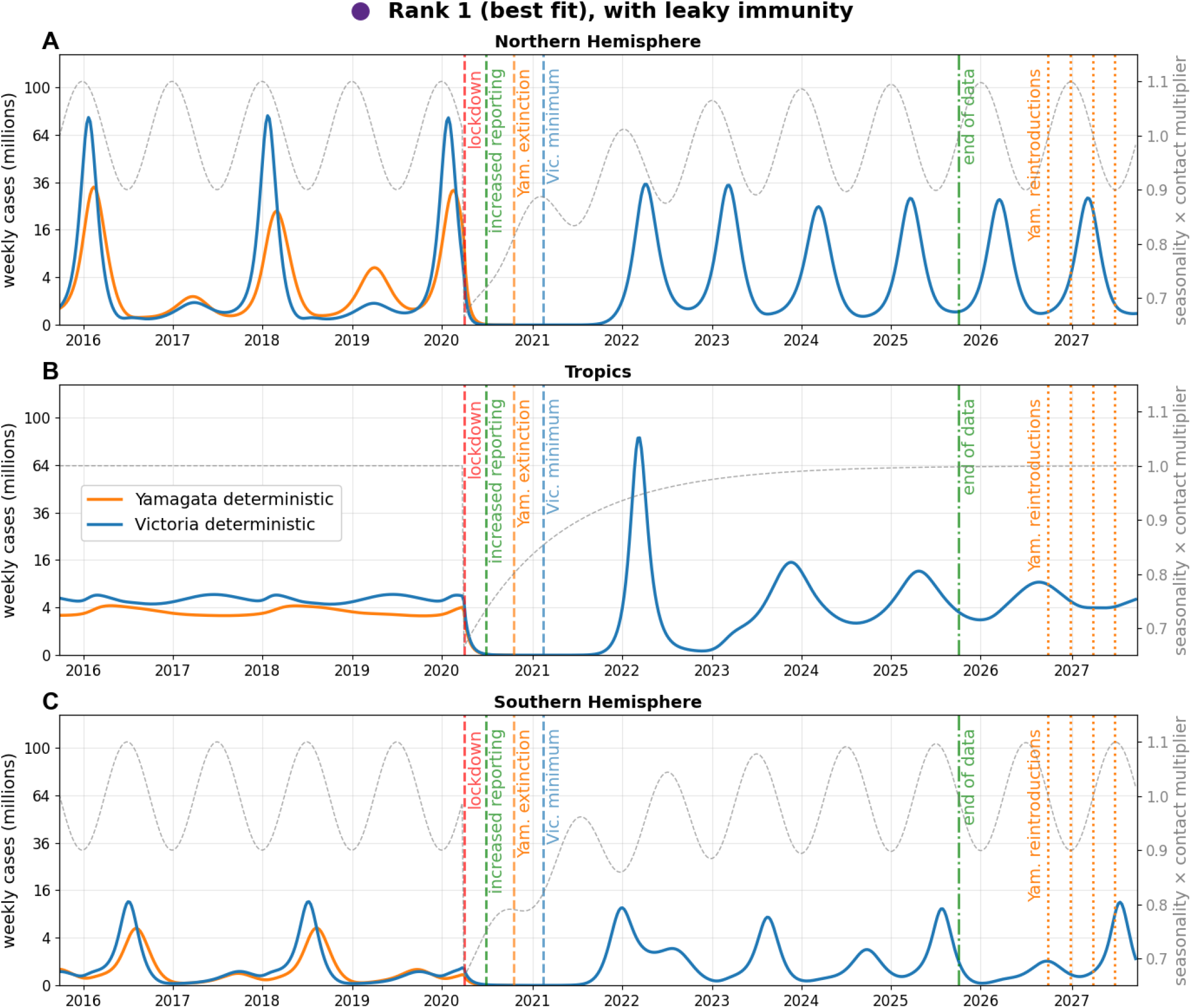
Model outcome for the best-fitting sample (Fig. 3 of the main text, same parameter values) with leaky instead of all-or-none cross-immunity. The dashed lines mark the extinction of Yamagata (orange) and the minimum global prevalence of Victoria (blue) in the leaky model.

**Fig. S13.**
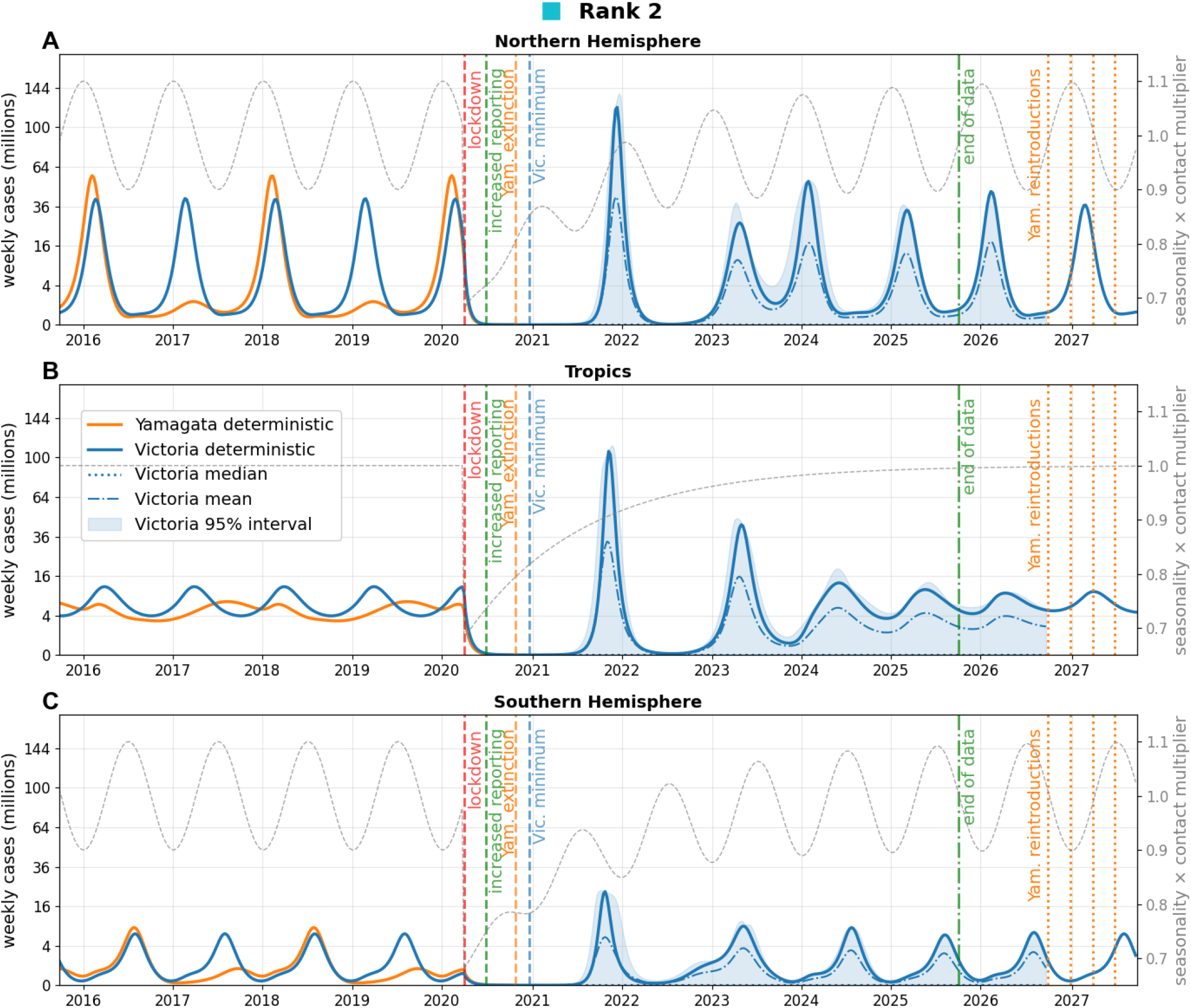
As Fig. 3 of the main text, but for the biologically-filtered sample with the second-highest likelihood (rank 2 in Table S1). The parameter values are *R*_0,*V*_ = 3.22, *R*_0,*Y*_ = 1.99, *T*_*V*_ = 7.10, *T*_*Y*_ = 6.86, *σ*_*V Y*_ = 0.01, *σ*_*Y V*_ = 0.00, *ϕ* = 8.9, *m* = 0.009, *c*_0_ = 0.32, *τ* = 476 (keys and units in Table 1 of the main text). The pre- and post-pandemic reporting rates that maximize the likelihood are *ρ*_*−*_ = 0.021% and *ρ*_+_ = 0.034%. The orange dotted lines represent the hypothetical dates of Yamagata re-introduction (see Fig. 4 of the main text). The blue dashed line marks the minimum global prevalence of Victoria according to the deterministic model.

**Fig. S14.**
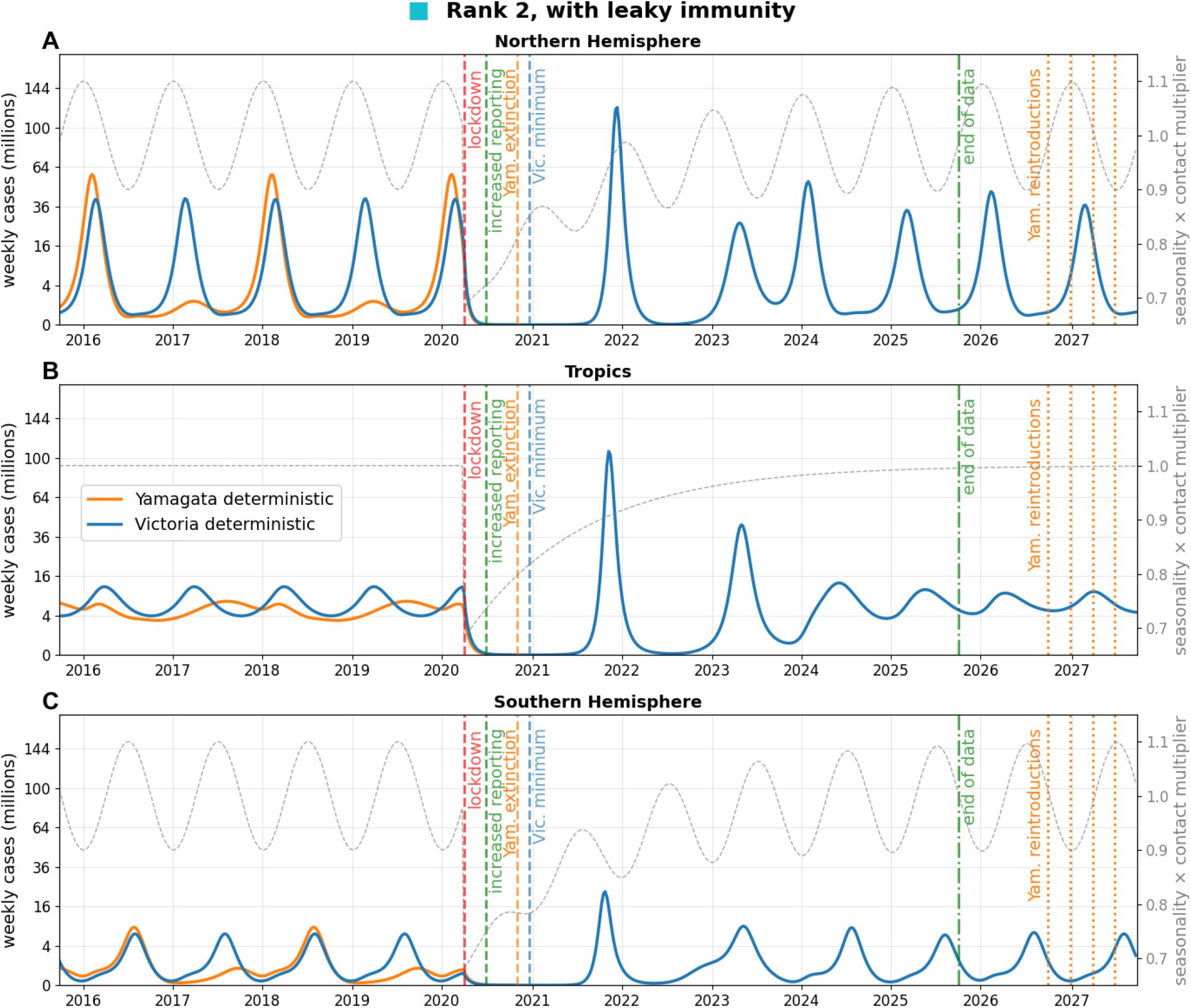
As Fig. S12, for the sample with the second-highest likelihood (rank 2 in Table S1), whose outcome with all-or-none cross-immunity is shown in Fig. S13.

**Fig. S15.**
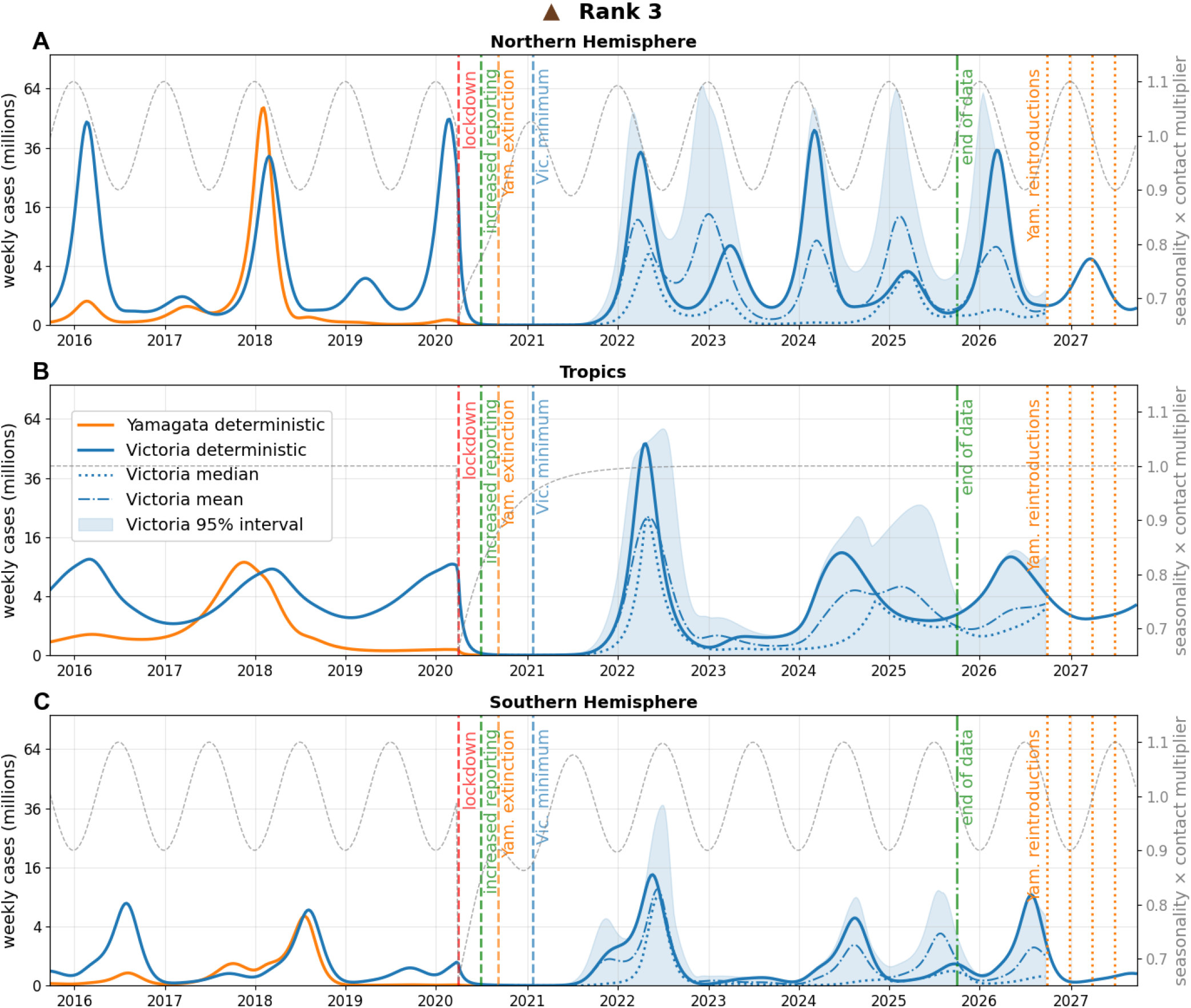
As Fig. S13, for the sample with the third-highest likelihood (rank 3 in Table S1). The parameter values are *R*_0,*V*_ = 1.61, *R*_0,*Y*_ = 1.55, *T*_*V*_ = 6.08, *T*_*Y*_ = 4.87, *σ*_*V Y*_ = 0.82, *σ*_*Y V*_ = 0.14, *ϕ* = 3.5, *m* = 0.009, *c*_0_ = 0.35, *τ* = 154 (keys and units in Table 1 of the main text). The pre- and post-pandemic reporting rates that maximize the likelihood are *ρ*_*−*_ = 0.039% and *ρ*_+_ = 0.061%.

**Fig. S16.**
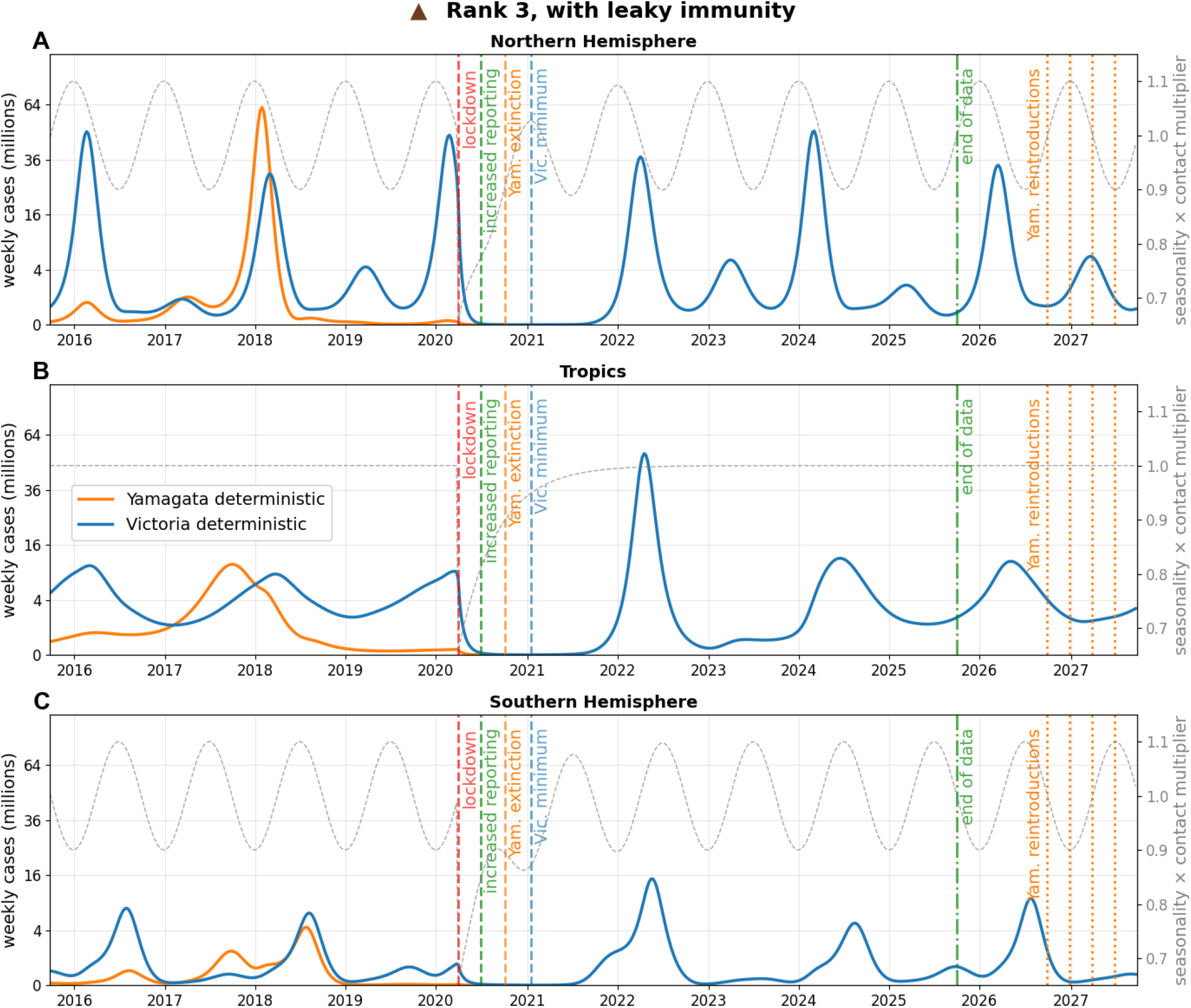
As Fig. S12, for the sample with the third-highest likelihood (rank 3 in Table S1), whose outcome with all-or-none cross-immunity is shown in Fig. S15.

**Fig. S17.**
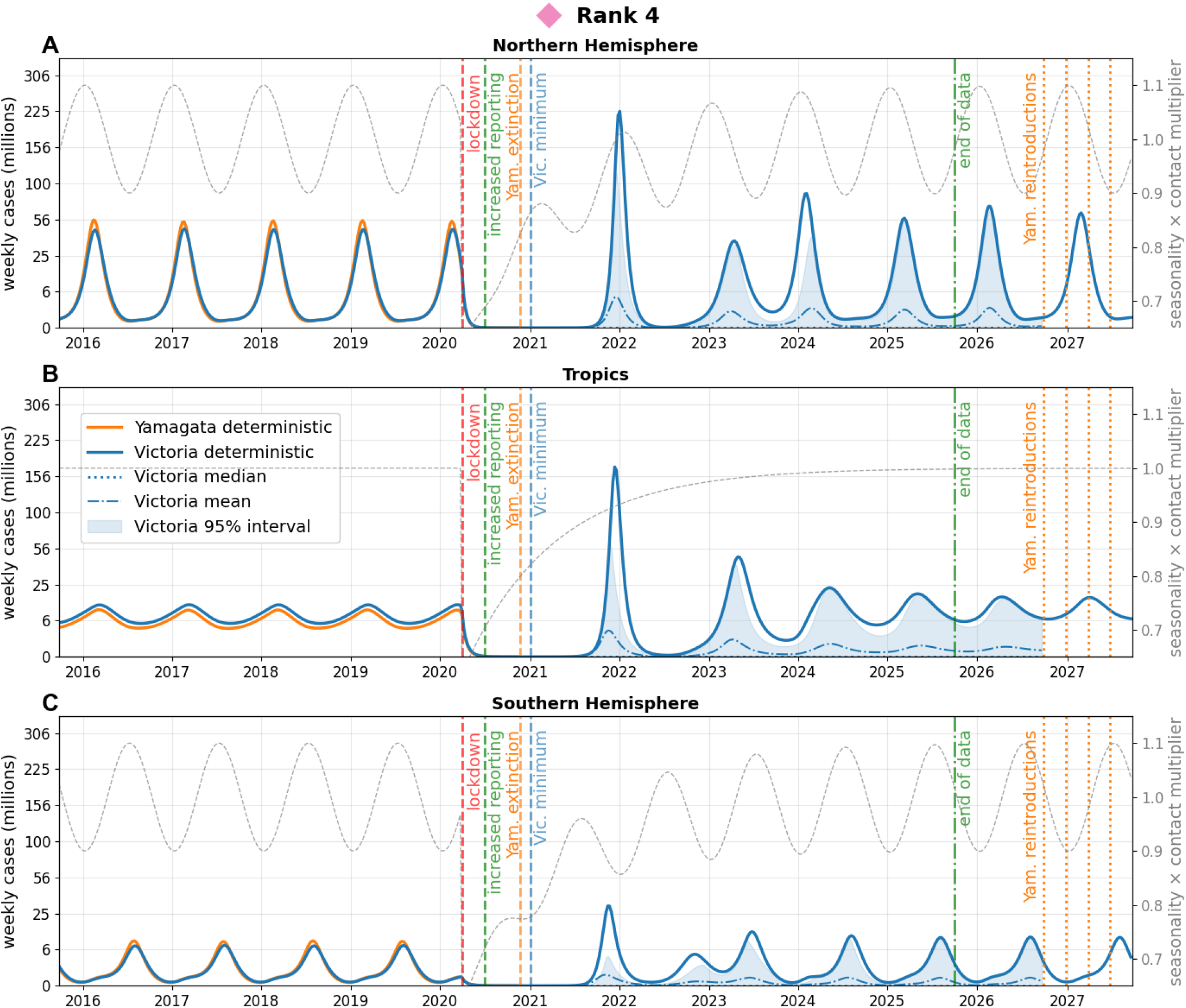
As Fig. S13, for the sample with the fourth-highest likelihood (rank 4 in Table S1). The parameter values are *R*_0,*V*_ = 2.17, *R*_0,*Y*_ = 1.50, *T*_*V*_ = 3.43, *T*_*Y*_ = 1.28, *σ*_*V Y*_ = 0.90, *σ*_*Y V*_ = 0.54, *ϕ* = 15.2, *m* = 0.008, *c*_0_ = 0.38, *τ* = 371 (keys and units in Table 1 of the main text). The pre- and post-pandemic reporting rates that maximize the likelihood are *ρ*_*−*_ = 0.017% and *ρ*_+_ = 0.022%.

**Fig. S18.**
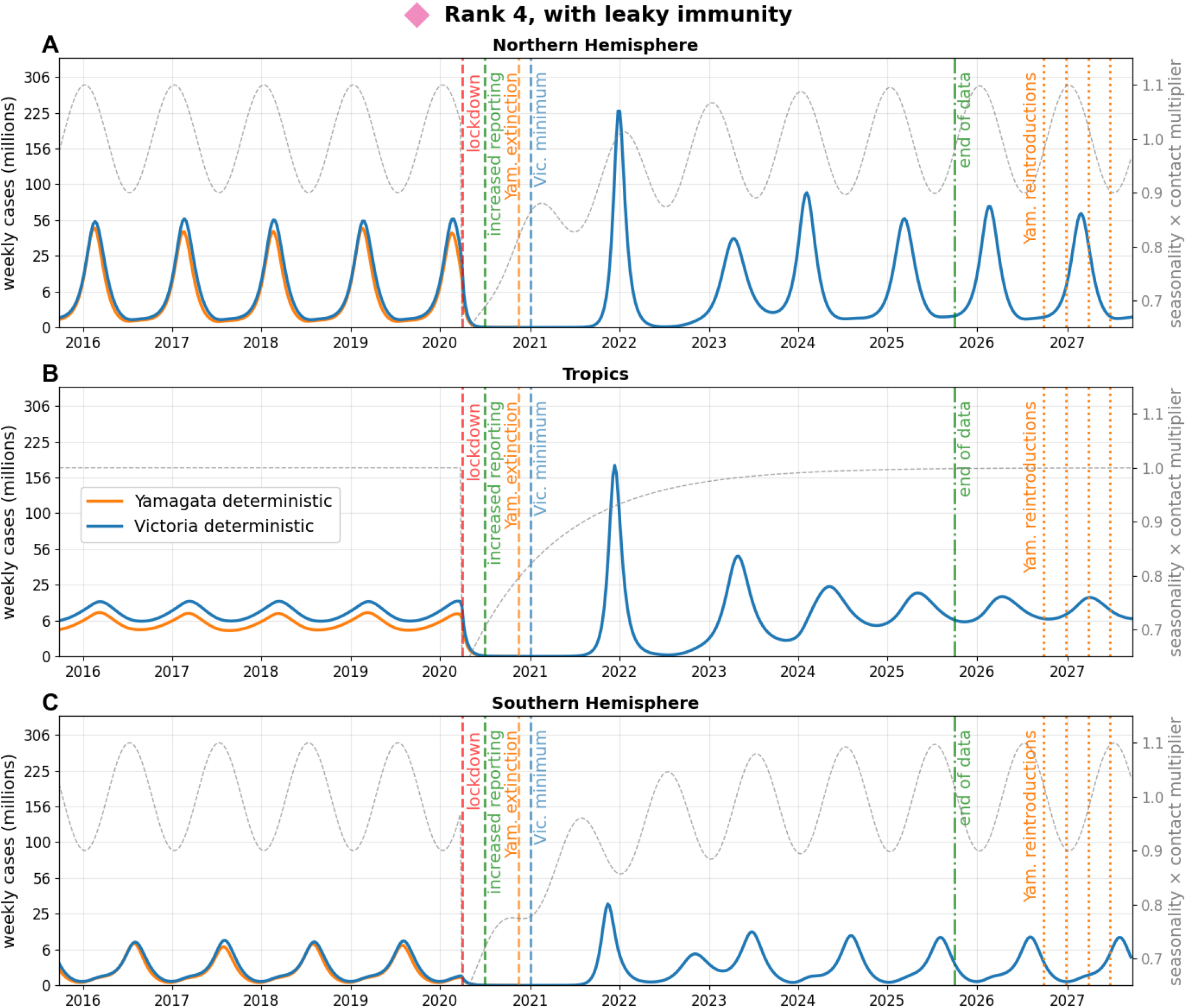
As Fig. S12, for the sample with the fourth-highest likelihood (rank 4 in Table S1), whose outcome with all-or-none cross-immunity is shown in Fig. S17.

**Fig. S19.**
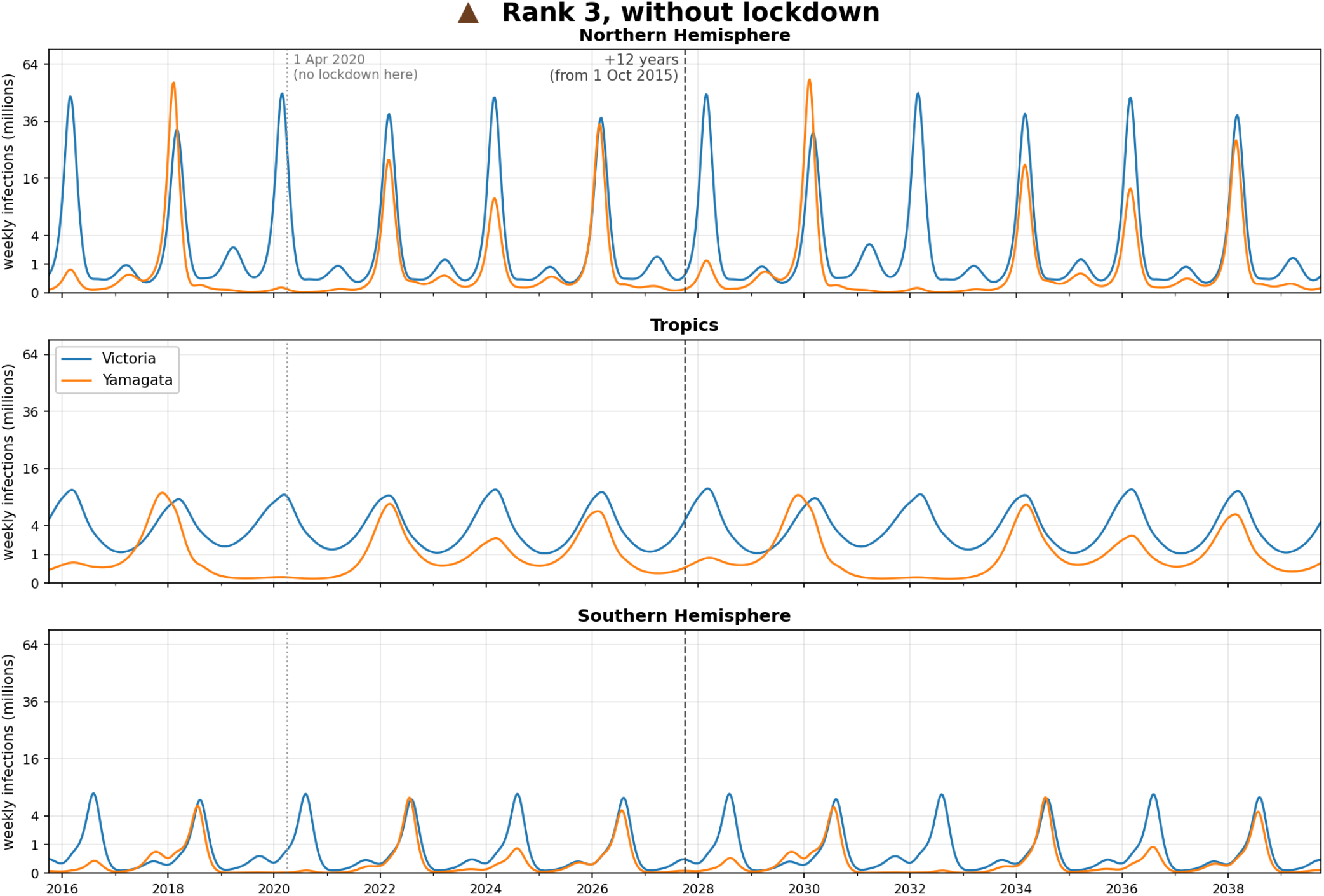
The sample with the third-highest likelihood (model output in Fig. S15), but without the lockdown. Before 2020 this sample has a large Yamagata season in the NH in 2017–18 followed by three years of near-inexistent Yamagata outbreaks. Without the lockdown the pattern nearly repeats 12 years later (dashed line): a large Yamagata season in 2029–30 followed by three near-empty ones. The dotted line marks 1 April 2020, when the lockdown would have started.

**Fig. S20.**
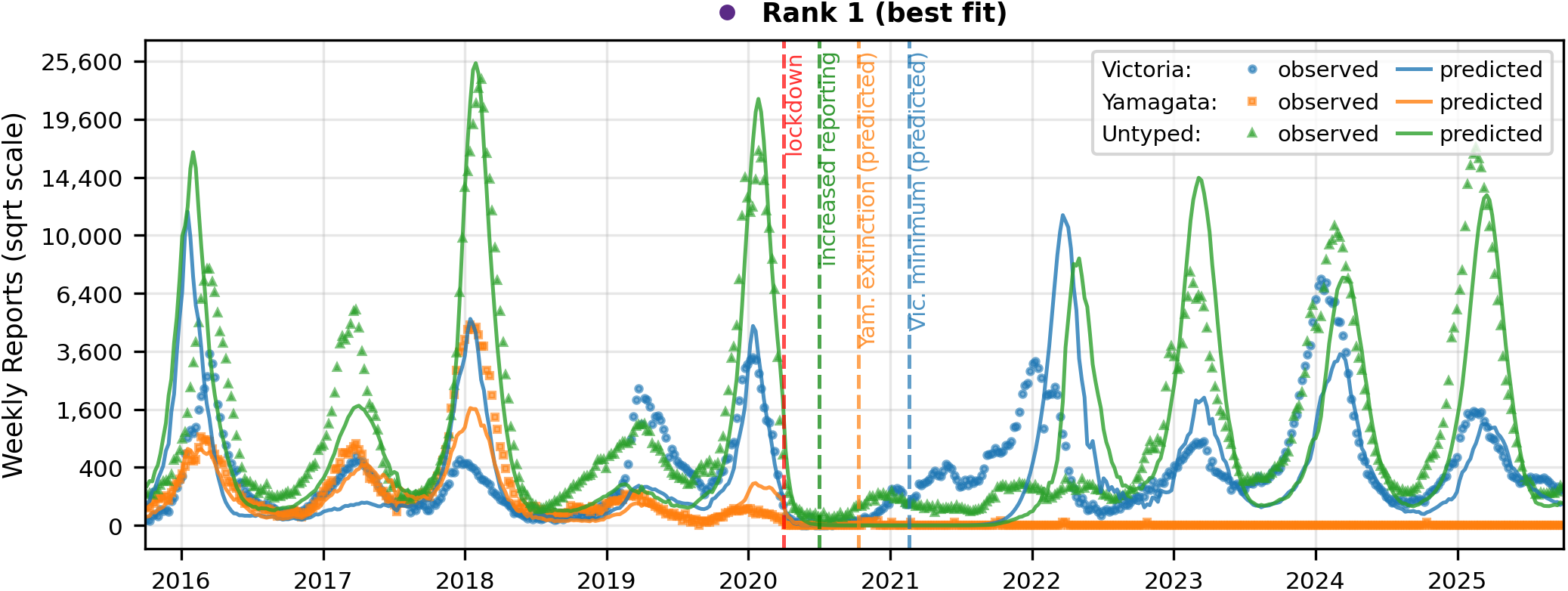
The best-fitting sample (Fig. 3 of the main text) against the weekly influenza B reports in the Northern Hemisphere FluNet data (Fig. 1 of the main text). Markers: reported Victoria, Yamagata and untyped (i.e., lineage not determined) influenza B infections. Lines: the weekly reports expected from the model, using the reporting rates *ρ*_*−*_ = 0.031% (until 30 June 2020) and *ρ*_+_ = 0.046% (from 1 July 2020).

**Fig. S21.**
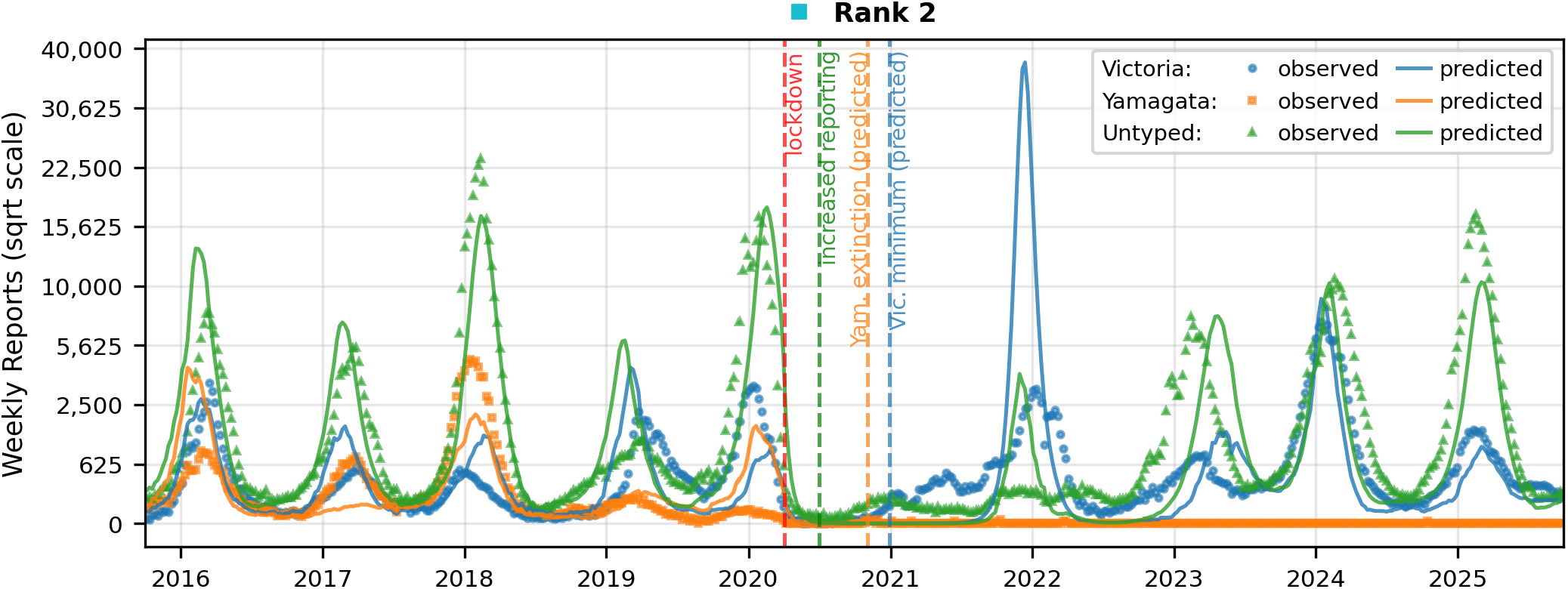
As Fig. S20, for the sample with the second-highest likelihood (Fig. S13). The reporting rates are *ρ*_*−*_ = 0.021% and *ρ*_+_ = 0.034%.

**Fig. S22.**
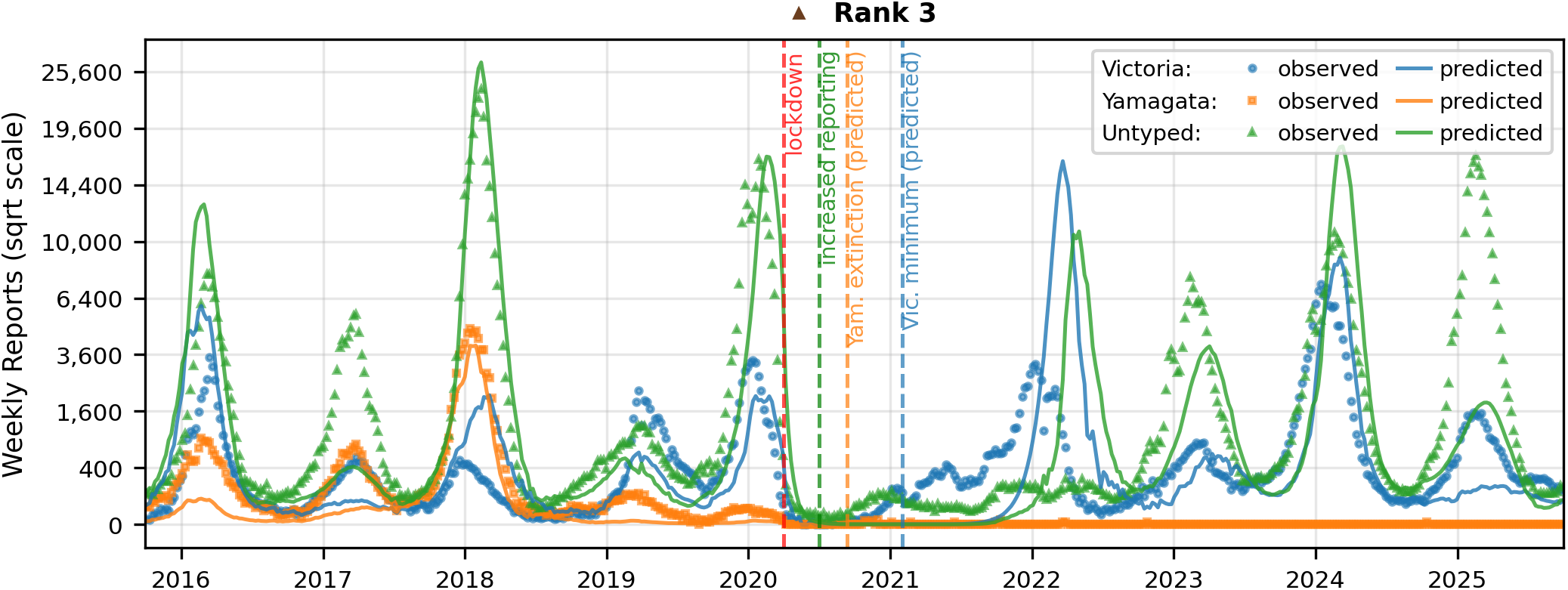
As Fig. S20, for the sample with the third-highest likelihood (Fig. S15). The reporting rates are *ρ*_*−*_ = 0.039% and *ρ*_+_ = 0.061%.

**Fig. S23.**
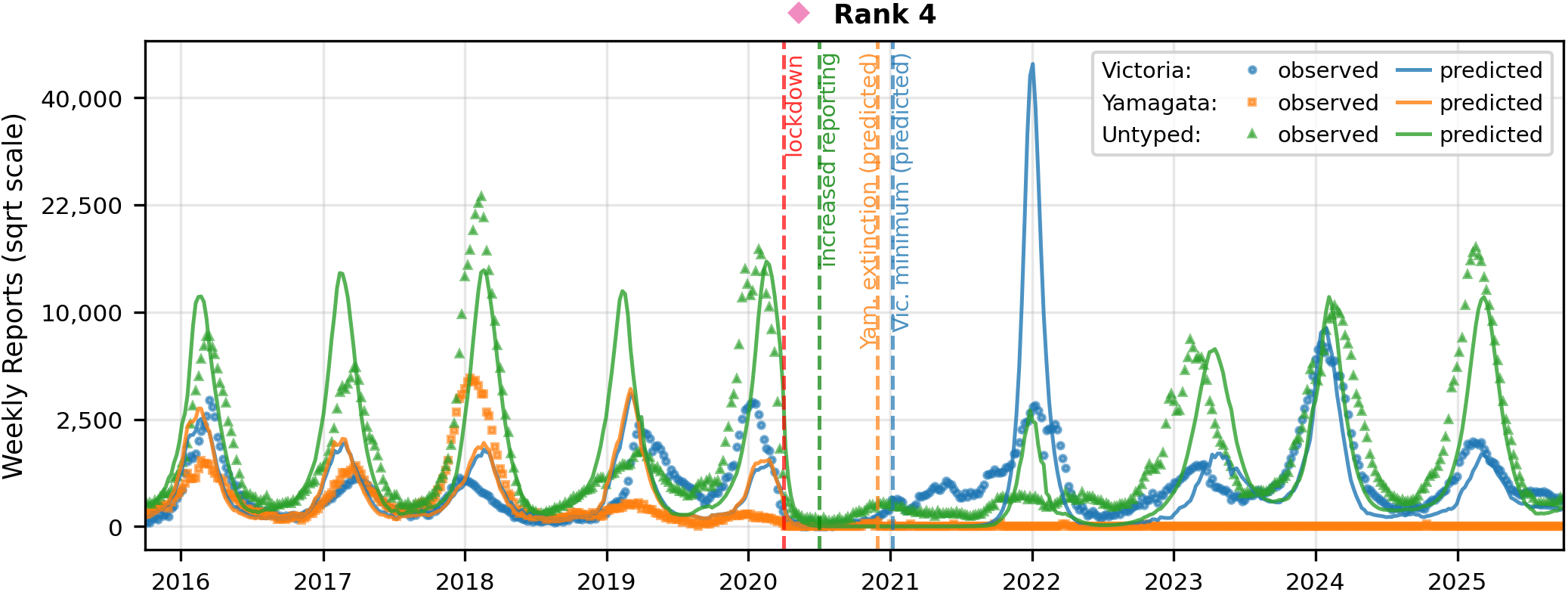
As Fig. S20, for the sample with the fourth-highest likelihood (Fig. S17). The reporting rates are *ρ*_*−*_ = 0.017% and *ρ*_+_ = 0.022%.

## Notes

### Competing Interest Statement

The authors have declared no competing interest.

### Summary of Updates

Modified force of infection, 1 million parameter samples re-evaluated with a new best-fitting sample.

